# Mapping DTP1,3 and MCV1 coverage and zero-dose prevalence in Nigeria: A spatiotemporal analysis (2000 – 2024)

**DOI:** 10.64898/2026.01.19.26344414

**Authors:** C. Edson Utazi, Mohamed Megheib, Iyanuloluwa D. Olowe, Somnath Chaudhuri, Natalia Tejedor-Garavito, George Mwinnyaa, Yoshito Kawakatsu, Danielle Boyda, Josh Lorin, Andrew J. Tatem

## Abstract

Spatially detailed estimates of childhood vaccination coverage are crucial to guide program design, targeting interventions, and evaluating progress within countries. In settings where substantial geographic inequities persist, high-resolution vaccination coverage and corresponding zero-dose maps can be helpful for understanding local patterns and informing strategies to reach underserved or missed populations. In this study, we produce annual 1×1 km and district level estimates of coverage for the first and third doses of diphtheria–tetanus–pertussis vaccine (DTP1 and DTP3) and the first dose of measles-containing vaccine (MCV1), as well as zero-dose prevalence, across Nigeria from 2000 to 2024. Our analyses draw on data from five Demographic and Health Surveys and two Multiple Indicator Cluster Surveys conducted between 2003 and 2024, alongside a suite of geospatial covariates. We fitted and evaluated Bayesian geostatistical models using the INLA-SPDE framework applied to cluster-level survey data. The resulting estimates highlight a persistent north–south divide in coverage, with consistently lower rates in the northern regions across the study period. Minimal gains were observed prior to 2015, followed by marked improvements that peaked in 2019 and declined slightly thereafter, with Jigawa and Yobe showing more sustained progress. We estimate that more than two million zero-dose children reside in Nigeria each year, with the highest burdens concentrated in the northeast, northwest, and parts of the south. These high-resolution outputs provide critical evidence to support subnational prioritization, strengthen routine immunization, and accelerate progress toward equitable vaccination coverage and disease elimination in Nigeria.

**What is already known on this topic:**

- Vaccination is one of the most successful public health interventions, saving millions of lives each year and making a major contribution to child survival.
- However, substantial gaps in vaccination coverage remain, particularly among children living in low- and middle-income countries (LMICs).
- Zero-dose and under-vaccinated children remain vulnerable to vaccine-preventable diseases and reaching them is crucial to prevent disease transmission and outbreaks.
- Reliable, current and spatially detailed evidence on coverage and the sizes and geographical distribution of zero-dose children is often unavailable in many LMICs, making the design of targeted interventions more challenging.

**What this study adds:**

- We employed Bayesian geostatistical modelling approaches to produce annual vaccination coverage and zero-dose estimates for Nigeria at 1×1 km resolution, the district and other administrative levels, covering the period from 2000 to 2024 (including predictions for 2025).
- Our results showed substantial heterogeneities and a persistent north-south divide in coverage. These also revealed that more than two million zero-dose children reside in Nigeria each year, with the highest burdens concentrated in the northeast, northwest, and parts of the south.

**How this study might affect research, practice or policy:**

- Our outputs can be triangulated with subnational administrative data for data quality assessments and other complementary datasets, such as travel time to the nearest health facility, to produce additional operationally relevant outputs.
- Using these outputs, program managers and policy makers can plan and implement targeted interventions to reach zero-dose and under-immunized children, hence accelerating progress towards Immunization Agenda 2030 goals in Nigeria.

## Introduction

Vaccination remains one of the most successful and cost-effective public health interventions [1]. Since its inception in 1974, the WHO Expanded Programme on Immunization (EPI) has been central to global infectious disease elimination and eradication efforts, averting an estimated 154 million deaths and making an unparalleled contribution to child survival [1, 2]. Despite substantial improvements in access to vaccination services many children - especially those living in low- and middle-income countries (LMICs) - continue to miss out on life-saving vaccines [3]. Reaching the ‘last mile’ populations, including those living in remote rural, conflict-affected settings, urban slums and highly mobile communities, remains a persistent challenge [4–6]. In 2024 alone, an estimated 19.9 million children were un- or under-vaccinated globally, of whom 14.3 million children were zero-dose - operationally defined as those who had not received the first dose of the diphtheria-tetanus-pertussis vaccine (DTP1) [3]. Geographic barriers, socioeconomic disadvantage, weak health infrastructure, insecurity and other forms of multiple deprivation continue to impede equitable access to vaccination [7]. These challenges highlight the need for targeted and innovative strategies to ensure that no one is left behind, as emphasized in global health policy frameworks such as the Sustainable Development Goals and the Immunization Agenda 2030 [8, 9].

Vaccination coverage is a fundamental metric for assessing the performance and effectiveness of immunization programmes. It provides essential information for prioritizing vulnerable populations, identifying missed communities, and diagnosing areas of weak programme performance, such as high dropout rates between successive doses in multi-dose vaccine series [10, 11]. Coverage is typically estimated using administrative data or household surveys. Although substantial efforts are underway to strengthen administrative data systems, many LMICs continue to face challenges in generating administrative data of sufficient accuracy and completeness for programmatic decision-making [12, 13]. Consequently, household surveys remain the gold-standard source of vaccination coverage data in these settings and can be triangulated with administrative records to enhance data quality and yield deeper insights into coverage patterns and trends [10]. However, because household surveys are expensive and logistically intensive, they are often designed only to provide estimates at national and first administrative levels, limiting their utility for planning targeted interventions at lower levels of the health system. Over the past decade, increasing attention has been directed toward overcoming this limitation, particularly through the development of modelling approaches capable of estimating coverage at small area scales using survey data [4, 14–21]. A range of methodological approaches - from geostatistical to machine learning and hybrid techniques - has been developed to support this work [22]. These methods enable estimation at finer spatial scales, such as districts or second administrative units where immunization programmes are implemented. Importantly, grid-level estimates are not constrained by changing administrative boundaries and can be readily integrated with gridded population estimates and ancillary datasets (e.g., travel time to the nearest health facility [23]), thereby supporting more nuanced analyses and broader programmatic utility.

Nigeria remains a priority country for the global immunization community, particularly in the effort to reach zero-dose and under-vaccinated children and underserved communities [24]. In 2024, Nigeria was among the top ten countries with the largest number of zero-dose children globally, recording the highest estimated numbers of both DTP-unvaccinated (2.1 million) and MCV-unvaccinated (3.1 million) children [3]. WHO and UNICEF estimates of national immunization coverage (WUENIC) for 2024 place DTP1, DTP3, and MCV1 coverage in Nigeria at 71%, 67%, and 57%, respectively, with MCV1 coverage not exceeding 60.3% at any point since 2010. These persistently low levels partly explain why measles remains endemic, with 10,948 cases reported in 2024 and 20,052 cases already reported in 2025 [25]. Despite these challenges, Nigeria remains committed to achieving its immunization and disease elimination goals, including measles and rubella elimination by 2030 [26]. The national immunization programme is implementing a range of targeted routine immunization strategies and conducting regular vaccination campaigns to accelerate progress. In 2022, a joint Gavi–WorldPop workshop with the National Primary Health Care Development Agency (NPHCDA) identified and prioritized 100 local government areas for intensified zero-dose interventions [27]. More recently, Nigeria launched the first phase of an integrated measles–rubella and polio vaccination campaign across 21 states (including the FCT) in 2025, with the second phase scheduled to begin in January 2026 across the remaining 16 southern states [28]. To complement these efforts, the country requires tools that can more precisely identify underserved populations, monitor subnational coverage trends, and support the effective planning of equitable vaccination strategies.

Since 2018, the VaxPop project at WorldPop, has undertaken various analyses to produce high-resolution maps of vaccination coverage [14, 17, 18, 29, 30{Utazi, 2025 #321]}. So far, our country-specific or multi-country geospatial analyses of vaccination coverage have typically been tied to individual surveys such as DHS, MICS or PCCS. Whilst these analyses provide highly accurate and timely outputs for immediate programmatic actions targeting vulnerable populations, there is substantial value in producing consistent subnational estimates of key vaccination coverage indicators over extended time periods. Such spatiotemporal estimates can support localised tracking of coverage trends, comparisons across birth cohorts, assessments of the impact of interventions (e.g., campaigns) and identification of persistent geographic patterns in the distribution of zero-dose and under-vaccinated populations. Motivated by these needs, the current analysis therefore aims to provide annual estimates of DTP1,3 and MCV1, as well as the dropout rates between DTP1 and DTP3, for children aged under one year of age (birth cohort year) at 1×1 km resolution and the district (or second administrative) level from 2000 to 2024. By adopting the birth-cohort year approach, our estimates can be readily integrated with administrative data, either for comparative assessments or to generate hybrid estimates of coverage that can leverage the strengths of both data sources.

## Methods

### Data

#### Vaccination coverage and measles campaign data

Data on DTP1, DTP3, and MCV1 coverage were obtained from seven nationally representative, geolocated household surveys conducted in Nigeria within the study period. These comprised the 2003, 2008, 2013, 2018, and 2024 Demographic and Health Surveys (DHS) and the 2016/17 and 2021 Multiple Indicator Cluster Surveys (MICS) [31, 32]. All surveys used stratified, multi-stage cluster sampling designs, with stratification achieved by dividing each of the 37 first-level administrative units (the 36 states and the Federal Capital Territory) into urban and rural areas. Within each stratum, clusters or enumeration areas were selected in the first stage and households within clusters in the second stage.

From each survey, we extracted information on eligible children’s ages, vaccination status, survey cluster coordinates, and relevant survey design variables. Vaccination status for each vaccine–dose combination was determined using both home-based records (HBRs) and caregiver recall; thus, our analysis focuses on crude coverage estimation. Each child was first assigned to one of four age groups based on their age at interview: 12–23, 24–35, 36–47, and 48–59 months. Using a birth-cohort year approach, we then assigned each age group to the calendar year in which the children would have been aged 0–11 months (i.e., their birth-cohort year). For example, the four age groups in the 2008 DHS were mapped to the 2007, 2006, 2005, and 2004 birth cohorts. Because vaccination data in the 2018 DHS, 2021 MICS, and 2024 DHS were collected only for children under 36 months, we were able to form only two birth-cohort groups from these surveys. For the 2016/17 MICS, we relied on pre-processed data from an earlier study [30] and included only children aged 12–23 months (the 2015 birth cohort). Details of the included birth cohorts and their assigned years are provided in Supplementary Table 1.

The birth-cohort year approach offers several advantages: (i) it increases the volume and temporal coverage of input data; (ii) it facilitates comparisons and integration with administrative coverage estimates; and (iii) it enables programmatically relevant attribution of gaps in coverage to specific birth cohorts. However, the approach assumes limited spatio-temporal influence of catch-up vaccination, migration, and mortality [16]. These assumptions, along with the potential for increased recall bias when using older cohorts from earlier surveys, have been examined elsewhere [16]. Additionally, our MCV1 estimates do not distinguish between doses received through routine services versus supplementary immunization activities (SIAs). We therefore assess the influence of SIAs on MCV1 coverage through comparisons with DTP3, consistent with previous work [18].

For each survey, birth-cohort year, and vaccine–dose combination, we aggregated child-level data to the survey-cluster level to derive the number of eligible children, the number vaccinated, and empirical cluster-level coverage. These cluster-level coverage data are shown in Supplementary Figures 1 – 3.

To support interpretation of MCV1 patterns, particularly in relation to DTP3, we also extracted data from WHO [33] on completed measles vaccination campaigns or SIAs conducted in Nigeria from 2005 to 2024. These are summarised in Supplementary Table 2.

#### Geospatial covariate data

Building on previous work [14, 17, 18, 30], we assembled a set of geospatial covariates for inclusion in the analysis. These comprised variables derived from DHS surveys - household wealth, maternal education, skilled birth attendance, ownership of a health card, proportion not stunted and access to media - as well as externally sourced covariates, including distance to urban areas, enhanced vegetation index, average maximum temperature, precipitation, and malaria prevalence. Detailed definitions and data sources for all variables are provided in Supplementary Tables 3 and 4. All selected covariates were temporally varying and were chosen based on their direct or proximate associations with vaccination coverage and their use in previous geospatial modelling studies [20, 34–36].

For the DHS-derived variables, we extracted cluster-level data from each survey using the definitions in Supplementary Table 3. These extracted values were matched to the calendar (birth-cohort) year corresponding to the 12–23-month age group from each survey. For example, data extracted from the 2018 DHS were assigned to the 2017 birth-cohort year. To generate annual, continuous surfaces for these variables, we fitted a simplified version of the model described in Equation (1), omitting covariates and relying solely on the spatio-temporal covariance structure to interpolate values across years.

Externally sourced geospatial covariates were processed and harmonised to a 1 x 1 km spatial resolution following procedures described in earlier work [14, 18]. Consistent with prior studies [14, 18], we extracted: (i) cluster-level values of DHS-derived covariates for all birth cohorts older than 12–23 months from the interpolated raster layers, and (ii) cluster-level values of the externally sourced geospatial covariates for all years in which survey input data were available (Supplementary Table 1).

#### Gridded population and administrative boundary data

Gridded population estimates for children aged under one year from 2000 to 2024 were obtained from WorldPop [37]. These estimates were adjusted to align with annual United Nations World Population Prospects totals [38], except for 2024, for which adjusted estimates were not available at the time of analysis. We used these population data in two ways: (i) to derive population-weighted administrative-level vaccination coverage estimates from the 1 x1 km grid-level predictions, and (ii) to calculate the annual numbers of zero-dose children. Administrative boundary data used in the analysis were sourced from GADM (www.gadm.org).

### Model fitting, prediction and validation

We adopted a Bayesian spatiotemporal geostatistical modelling framework to model and predict vaccination coverage at 1×1 km resolution. Letting *Y* (***s***_*i*_, *t*) denote the number of vaccinated children in birth-cohort year *t* and survey location ***s***_*i*_ (with known latitude and longitude coordinates), *m* (***s***_*i*_, *t*) the corresponding number of children sampled in the survey, *p* (***s***_*i*_, *t*) (0 ≤ *p* (***s***_*i*_, *t*) ≤ 1), and, the true unobserved vaccination coverage. The model can be expressed as:

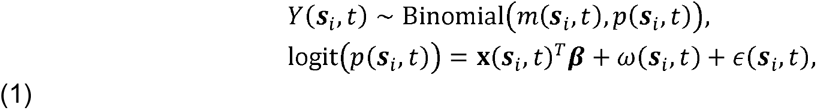

where **x** (***s***_*i*_, *t*) is a *p* × 1 vector of covariates and ***β*** the corresponding regression parameters, *ω* (***s***_*i*_, *t*) is a spatiotemporal random effect and *ϵ* (***s***_*i*_, *t*) is an identically and independently distributed Gaussian random effect with variance, 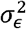, i.e., 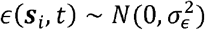. We model *ω* (***s***_*i*_, *t*) as a zero-mean spatiotemporal Gaussian process with a separable covariance structure [39–41] given by Σ_*ω*_ = Σ_*s*_ ⨂ Σ _*t*_, where ⨂ denotes the Kronecker product of two matrices. We assume Σ _*s*_ to follow the Matérn covariance structure [42] given by

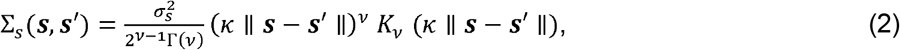

where ‖ · ‖ denotes the Euclidean distance, 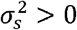 is a marginal variance parameter, *κ* is a scaling parameter related to the range 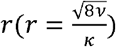 – the distance at which spatial correlation is close to 0.1, and *K*_*v*_ is the modified Bessel function of the second kind and order *v* > 0. For identifiability reasons, we set the smoothness parameter *v* = 1, see [43]. We further assume a first-order autoregressive (AR(1)) covariance structure for Σ_*t*_, such that 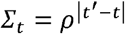 where *ρ* (− 1 < *ρ* < 1) is an autoregressive parameter governing the temporal dynamics in *ω* (***s***_*i*_, *t*) We note that it is possible to specify non-separable covariance functions for *ω* (***s***_*i*_, *t*) (see, e.g., Sahu [41]) but these are more computationally intensive to fit.

To complete the Bayesian specification, we placed a non-informative prior on the regression coefficients: ***β*** ~ *N* (**0**, 10^−3^ *I*). We further placed penalized complexity priors on *ρ,r, σ*_*ϵ*_and *σ*_*s*_ such that *p* (*ρ* > 0) = 0.9, *p* (*r* < *r*_0_) = 0.01, *p* (*σ*_*ϵ*_ >1) = 0.01, and *p* (*σ* > 3) = 0.01, where *r*_0_ is chosen to be 5% of the extent of the country in the north-south direction.

Model (1) was applied directly to mapping DTP1 and MCV1 coverage. To ensure that the estimates of DTP1 and DTP3 were internally consistent, i.e. *p*_*DTP*1_ (***s***, *t*) ≥ *p*_*DTP*3_ (***s***, *t*) for every ***s*** and *t*, we adopted the ratio-based approach implemented in Utazi et al [44]. This involved directly modelling the ratio of DTP1 and DTP3 *p*_*DTP*3/1_ (***s***, *t*), or DTP3/1, and then using the resulting posterior distribution and that of DTP1 to obtain estimates of DTP3.

In our model implementation, we note that although data are available for 18 out of 25 time points within the study period, when the model is fitted, predictions can be obtained for every single year within the study period, as well as for future time points.

The proposed model was implemented in a Bayesian framework using the integrated nested Laplace approximation – stochastic partial differential equation (INLA-SPDE) approach [43, 45, 46]. Using the fitted models, we obtained annual estimates of vaccination coverage covering the study period at 1×1 km resolution and for different administrative areas for each vaccine-dose combination. We also obtained grid level predictions for 2025. As in previous work [17], administrative level estimates were obtained as population-weighted averages of the grid level estimates falling within each area.

We assessed the out-of-sample predictive performance of the fitted models (for directly modelled indicators) at the cluster level by conducting a k-fold cross-validation exercise, in which we created the folds as random and spatially stratified partitions of the data, setting *k* = 10. We then used the following metrics to evaluate predictive performance: correlation between observed (*p* (***s***_*i*_, *t*)) and predicted 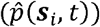 values, root mean square error, 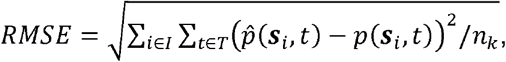, mean absolute error, 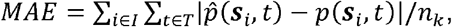, average bias, 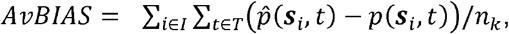, continuous ranked probability score, 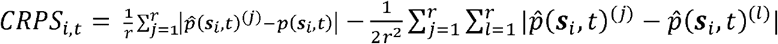 where *n*_*k*_ = | *I*| × |*T*| is the number of observations in the *k*th fold and *r* denotes the number of posterior samples, as the CRPS is averaged over all the locations within each fold and over all the *k* folds. All the other metrics are also averaged over the *k* folds. While these metrics are particularly useful for assessing the accuracy of the point estimates, the CRPS evaluates the accuracy of both the point and uncertainty estimates.

Additionally, for all the target indicators, we further examined the performance of the fitted models for each survey year by comparing the modelled estimates at both the regional and/or provincial (state) levels, depending on the representativeness of the survey, with direct survey estimates during the survey years.

## Results

### Parameter estimates and model validation

In Supplementary Tables 5 - 7, we report estimates of the parameters of the fitted models for DTP1, DTP3/1 and MCV1. Maternal education (+ve), skilled birth attendance (+ve), health card ownership (+ve) and malaria prevalence (−ve) were significantly associated with all three directly modelled indicators. Additionally, enhanced vegetation index (−ve) was significantly associated with DTP1 and MCV1, while distance to urban areas (−ve), maximum temperature (+ve) and access to media (+ve) were each significantly associated with DTP1, DTP3/1 and MCV1, respectively.

Th estimated values of the autoregressive parameters for the spatiotemporal process, *ω* (***s***_*i*_, *t*) ranged between 0.78 and 0.86, suggesting very strong temporal dependence in the model and slow changes over time. The estimated ranges were 228 km, 183 km and 287 km for DTP1, DTP3/1 and MCV1, indicating stronger, more localised spatial dependence in DTP3/1 unaccounted for by the covariates included in the model, compared to DTP1 and MCV1. In contrast, for DTP1 and MCV1, the spatiotemporal process, *ω* (***s***_*i*_, *t*) accounted for greater portion of the total residual variance, whereas for DTP3/1, the IID term *ϵ* (***s***_*i*_, *t*) had relatively greater variation in the model.

Our assessment of the predictive performance of the fitted models at the cluster level (Table 1) showed that vaccination coverage was reasonably well predicted, with out-of-sample correlations estimated to be 0.74, 0.67 and 0.47 for DTP1, MCV1 and DTP3/1, respectively under random cross-validation. The MAE values ranged between 19% (DTP1) and 26% (DTP3/1), while the AVGBIAS values indicate a tendency to slightly underestimate coverage on average in all the fitted models. Other validation metrics exhibited similar patterns, with the target indicators, DTP1 and MCV1, being better estimated than the intermediate, directly modelled ratio indicator, DTP3/1, further highlighting the difficulty in estimating the indicator. Also, as expected, predictive performance under random cross-validation was better than spatially stratified cross-validation in all cases.

**Table 1:** Results of cluster-level k-fold cross-validation to assess the out-of-sample predictive performance of the fitted models.

| Indicator | Correl. | RMSE | MAE | AVGBIAS | CRPS |
| --- | --- | --- | --- | --- | --- |
| Random k-fold |  |  |  |  |  |
| DTP1 | 0.743 | 0.251 | 0.191 | -0.012 | 0.143 |
| DTP3/1 | 0.473 | 0.318 | 0.263 | -0.039 | 0.167 |
| MCV1 | 0.667 | 0.281 | 0.223 | -0.010 | 0.171 |
| Stratified k-fold |  |  |  |  |  |
| DTP1 | 0.501 | 0.267 | 0.204 | -0.010 | 0.147 |
| DTP3/1 | 0.315 | 0.331 | 0.281 | -0.050 | 0.176 |
| MCV1 | 0.465 | 0.296 | 0.240 | -0.007 | 0.177 |

At both the regional and state levels, we observed strong correspondence between the modelled estimates and the direct survey estimates for DTP1,3 and MCV1across the survey years included in our study (Supplementary Figures 4 – 9). However, the correspondence was stronger for DTP1 and MCV1 than DTP3, especially at the state level where there is evidence of some underestimation of higher values of DTP3. This further corroborates that our modelling framework more robustly estimates directly modelled indicators.

### 1×1 km and aggregate estimates of DTP1,3 and MCV1 coverage at the national, state and local government area levels (2000 – 2024)

In Figures 1 – 2 and supplementary Figures 10 – 14, we present the 1×1km grid level estimates of vaccination coverage, associated uncertainties (95% credible interval widths) and absolute differences in coverage during the study period. Visualizations of these estimates are also available via the link: https://som-shinyapps.shinyapps.io/nga_vacn_explorer/. Figure 1 shows a characteristic north-south divide in DTP1 coverage, with persistent lower coverage areas in the north over the years indicating persistent challenges in accessing immunization services in these locations. The general trend in coverage is such that there were very little or no annual improvements in coverage between 2000 and 2015 (Figure 2). This is then followed by a gradual rise in coverage, appearing to peak in 2019 and then falling slightly in more recent years. Notably, from 2021, coverage appears to have fallen to pre-2015 levels in the northwest.

**Figure 1:**
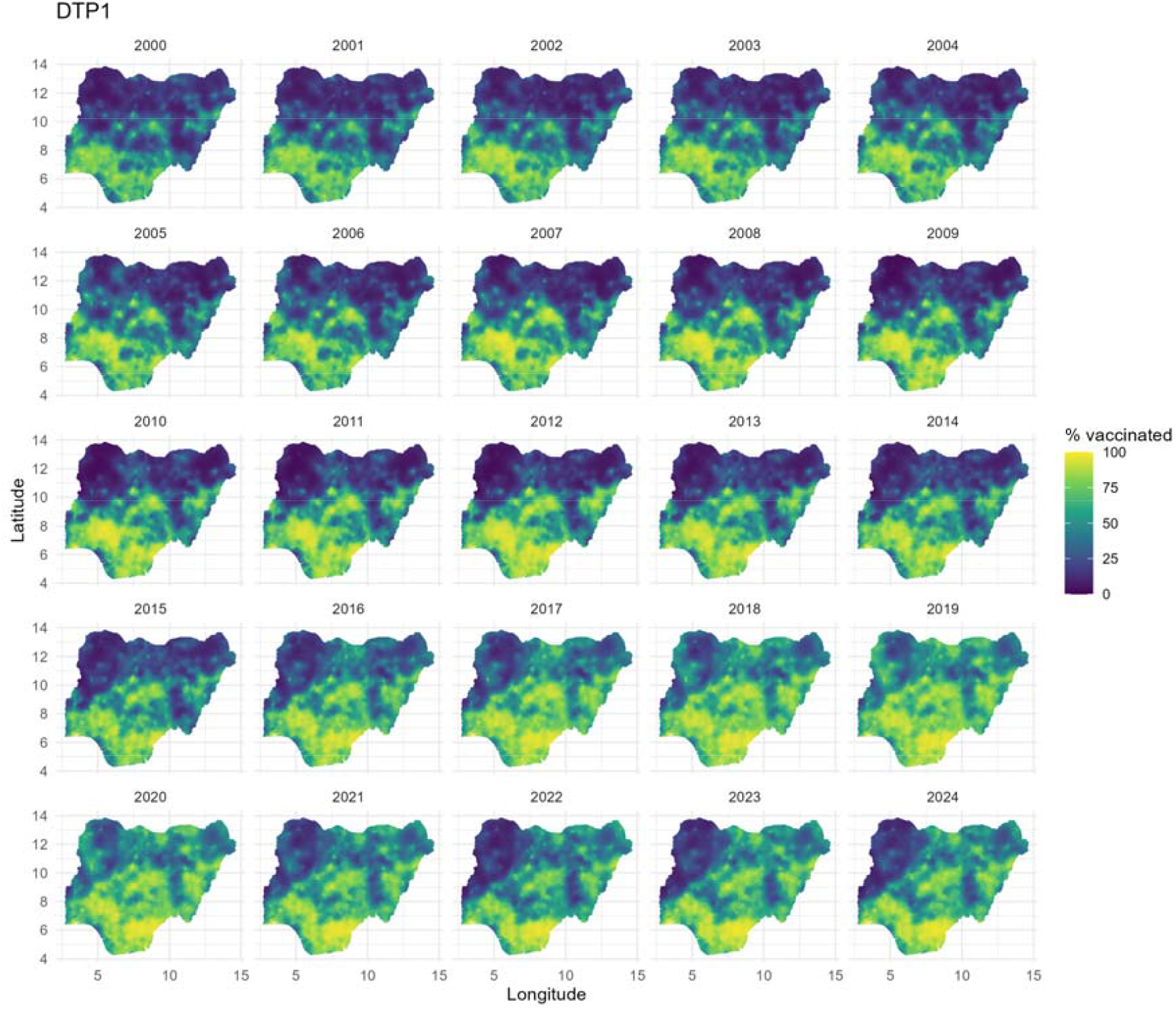
1×1 km estimates of DTP1 coverage for Nigeria from 2000 to 2024. These estimates relate to the birth cohort in each year. The associated uncertainties are displayed in supplementary Figure 12.

**Figure 2:**
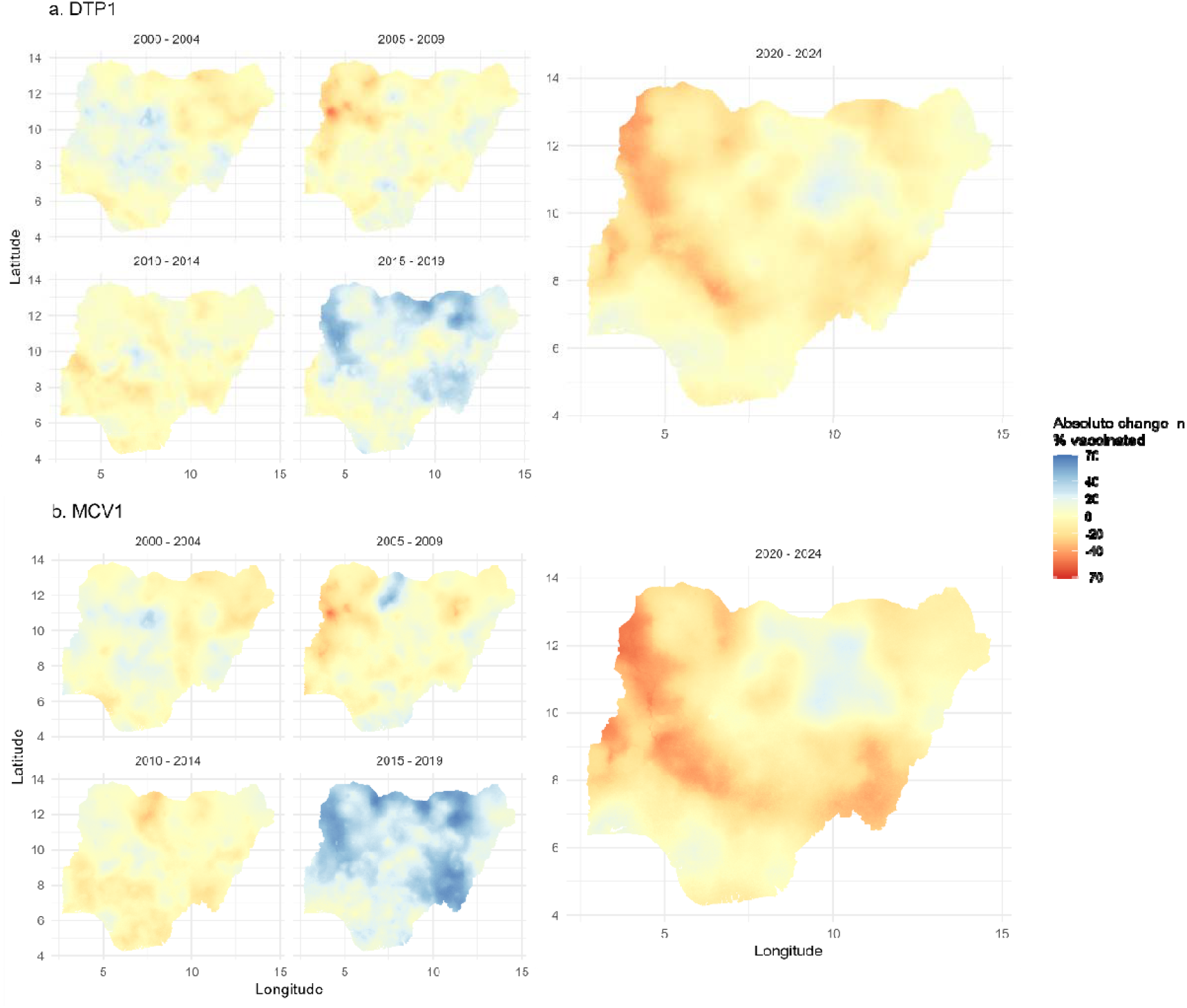
Absolute changes in (a) DTP1 and (b) MCV1 coverage in Nigeria between 2000 and 2024 at 1×1 km resolution. The coverage estimates used to calculate these changes relate to the birth cohort in each year. Positive values indicate increases in coverage within the stated period.

Broadly similar spatiotemporal patterns can also be observed in the maps of DTP3 and MCV1 coverage shown in supplementary Figures 10 and 11, except that the estimated coverage levels for both indicators are generally lower than those of DTP1. Also, for MCV1 coverage, Figure 2 reveals stronger improvements in coverage relative to DTP1 in 2019 and slightly worse regression in coverage in many areas between 2020 and 2024. Notably, areas with the most regression in MCV1 coverage in 2024 are concentrated in Kebbi, Niger, Kwara and Taraba states. Predictions for 2025 (supplementary Figure 22) show similar patterns in coverage as 2024.

The uncertainty maps shown in supplementary Figures 12 – 14 generally reveal higher uncertainties for DTP3 relative to DTP1 and MCV1. Again, this is likely because the latter indicators were modelled directly, while DTP3 was derived from the other modelled indicators. Also, the uncertainties are lower in areas where higher or lower coverage is predicted than areas where the coverage estimates are closer to 50% - an artefact of the binomial distribution used in model (1). Another important pattern seen in the uncertainty maps is that areas with sparse or no data generally have greater uncertainties than areas with a high density of cluster locations (see Supplementary Figures 1 – 3). As noted in previous work [47], the greater uncertainties observed at the grid level reduces greatly when the coverage estimates are aggregated to the area level, e.g. the local government area (LGA) and state levels (see Figure 3 and Supplementary Figures 15 and 16).

**Figure 3:**
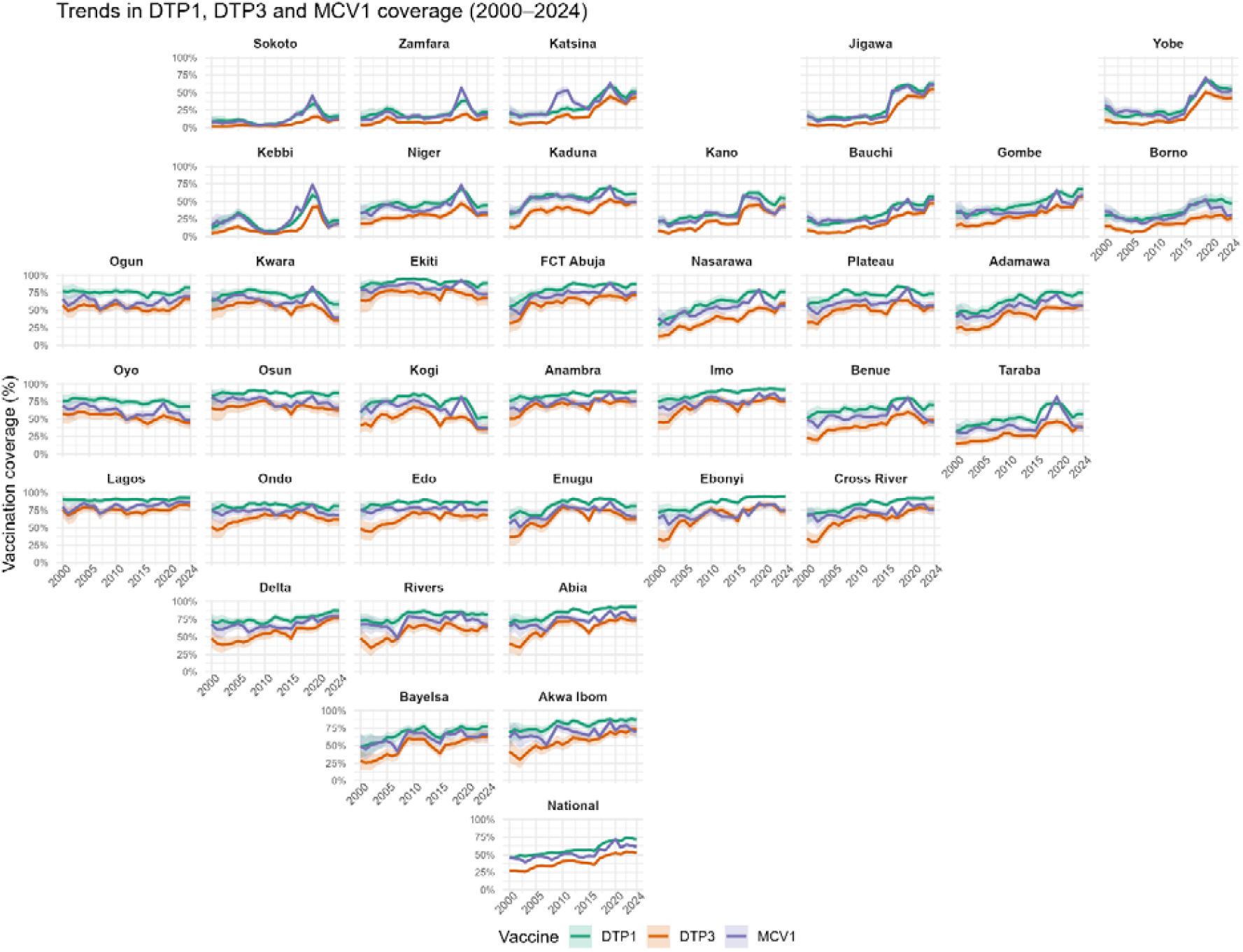
Trends in the estimates of DTP1,3 and MCV1 coverage at the state and national levels from 2000 to 2024 (year of birth). The plots are arranged to show the relative geographical positions of the states.

At the national level and in some states, an overall increasing trend in coverage is apparent for all three vaccine-dose combinations (see Figure 3; interactive visualizations of all administrative level estimates can be accessed via https://som-shinyapps.shinyapps.io/nga_vacn_explorer/). This appeared to gain some momentum after 2015 but has slowed down in more recent years. Notably, coverage remained below the 75% coverage mark throughout the study period for all three indicators at the national level. At the state level, some remarkable patterns are apparent. The sharp increase in coverage post-2015 is most pronounced in the northeast and northwest, particularly in Jigawa and Yobe states, both of which continue to maintain the improved coverage levels. Other low coverage states in both regions, such as Kebbi, Sokoto and Zamfara states, also experienced dramatic rises in coverage during the same period, but coverage levels have returned to pre-2015 levels in more recent years in these states. Overall, coverage seemed poorest in Sokoto and Zamfara States throughout the study period.

At the district or LGA level, example plots in Supplementary Figures 15 and 16 reveal patterns similar to the grid level estimates. Notably, however, the uncertainties in the estimates have reduced considerably, compared to those of the grid level estimates.

We investigate the patterns in the dropout rates between DTP1 and DTP3 at the LGA level as shown in supplementary Figure 17. The highest dropout rates were estimated in lower coverage areas in the northeast and northwest between 2000 and 2010. Thereafter, significant progress could be seen in the period from 2011 to 2024; however, areas of relatively higher dropout rates are still concentrated in lower coverage LGAs in Sokoto, Zamfara, Borno and Kwara states in more recent years.

As noted previously, our analysis of MCV1 coverage included doses received both through routine immunization and vaccination campaigns. Hence, by comparing estimates of DTP3 and MCV1 coverage as in previous work [18], we can investigate whether measles vaccination campaigns have boosted MCV1 coverage beyond routine immunization levels. We note that each of the birth cohorts included in our analysis from 2005 to 2023 would have participated in at least one campaign activity (see Supplementary Table 2). However, the effect of vaccination campaigns on MCV1 coverage appears most pronounced in the period from 2015 to 2019, especially in the northern regions (Supplementary Figure 18). Also, from 2009 to 2011, MCV1 coverage seems substantially higher in Katsina State. Notably, from 2021 to 2024, there was no noticeable effect of campaigns on MCV1 coverage. Some areas of relatively higher MCV1 coverage can be observed in the period from 2009 to 2002, but we are unable to attribute this to the impact of SIAs due to data unavailability. We discuss approaches for estimating the effect of campaigns on MCV1 coverage further in the discussion section.

### Identifying low coverage areas for DTP1 and MCV1

Consistent patterns in the occurrence of low coverage areas can often help inform targeted strategies and transitioning from national to subnational intervention programmes. Mapping the spatiotemporal distribution of coldspots of low coverage at operationally relevant scales is therefore of substantial programmatic importance. In Figure 4 and Supplementary Figures 19 and 20, we calculated and mapped the probabilities of having DTP1 and MCV1 coverage of 40% or less at the district level, respectively, to identify coldspots of low coverage. These probabilities also incorporate the uncertainties associated with the district level predictions, since these were calculated using the entire posterior distribution generated for each area.

**Figure 4:**
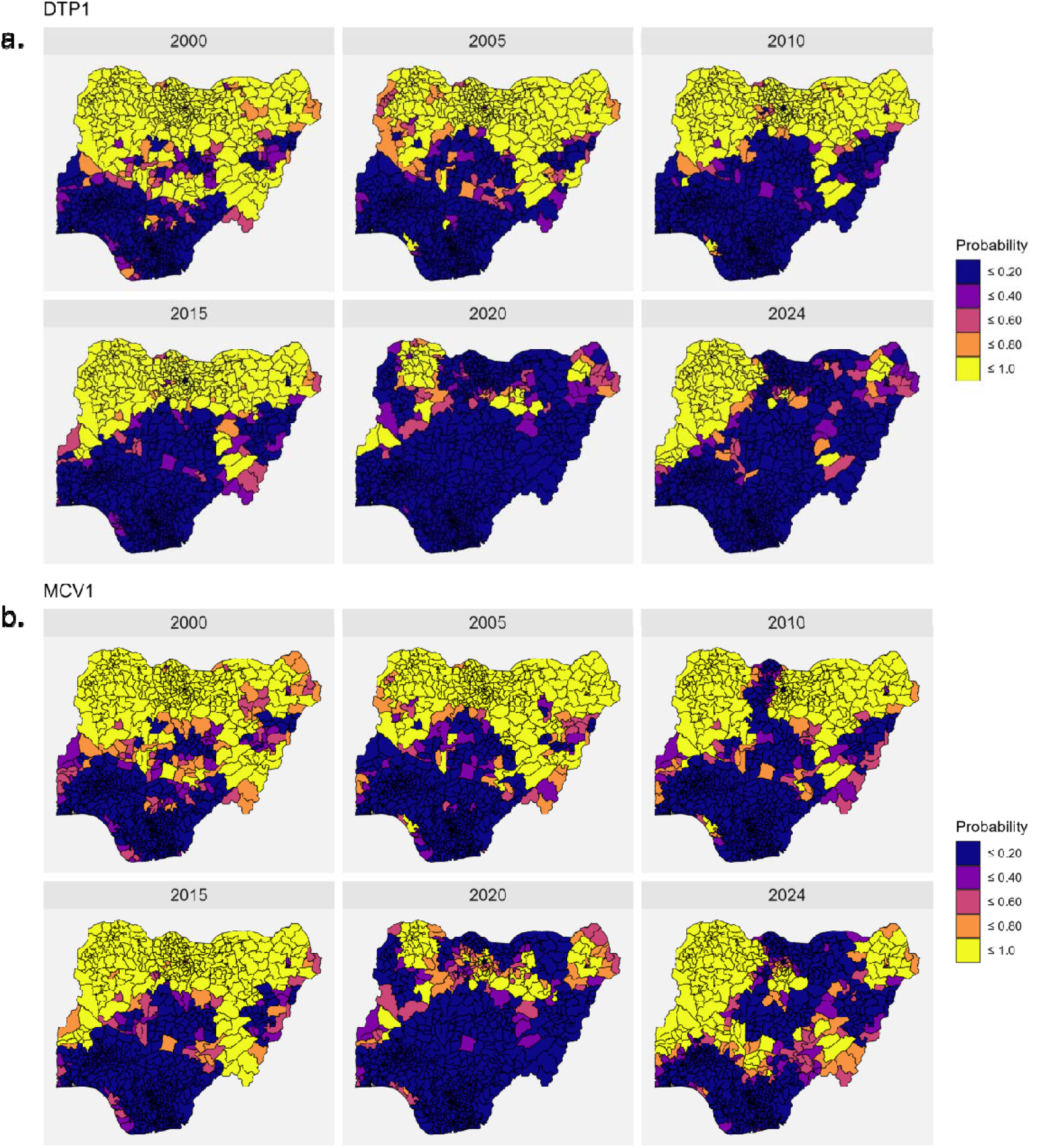
Probabilities of attaining an estimated MCV1 coverage of ≤40% at the local government area (LGA) level. Low coverage areas were defined as those with probabilities greater than 80%. The years shown correspond to the birth years of the cohorts included in the study.

For MCV1 (Figure 4b), the low coverage areas (those with probabilities > 0.8) are concentrated in the north, with a fairly consistent spatial distribution from 2000 to 2015. A reduction in both the geographic spread and numbers of low coverage districts is seen from 2016 to 2019 (supplementary Figure 20), with the smallest numbers of such low coverage areas observed between 2018 and 2020. However, from 2021, larger areas of low coverage with strong patterns in the western and northern parts of the country are visible. Notable in this period is also the concentration of coldspots of low coverage areas in the northeastern state of Borno and parts of Taraba state. We, however, note that due to the sparsity of input data in Borno state as a result of insecurity, the estimates produced for the state should be treated with caution. Similar patterns can also be observed in the low coverage areas estimated for DTP1 (Figure 4a and Supplementary Figure 19). However, the occurrence of coldspots of low coverage post 2020 is more concentrated in the northwestern areas of the country.

### Spatiotemporal trends in estimates of numbers of zero-dose children at the national, state and district levels

At the national level, estimates of numbers of (DTP) zero-dose children aged under one year old had been above 2 million annually from 2000 to 2024, except for 2019 for which an estimate of 1.96 million children was obtained (Figure 5a). Our analysis also revealed a slight upward trend in the numbers of zero-dose children from 2000 to 2016, as a result of increasing populations of under 1s during this period. This was then followed by some reductions, which appear to increase slightly towards 2022 and then stabilize in the later years. In particular, for 2023 and 2024, we obtained estimates of 2.21 million and 2.22 million zero-dose children, respectively.

**Figure 5:**
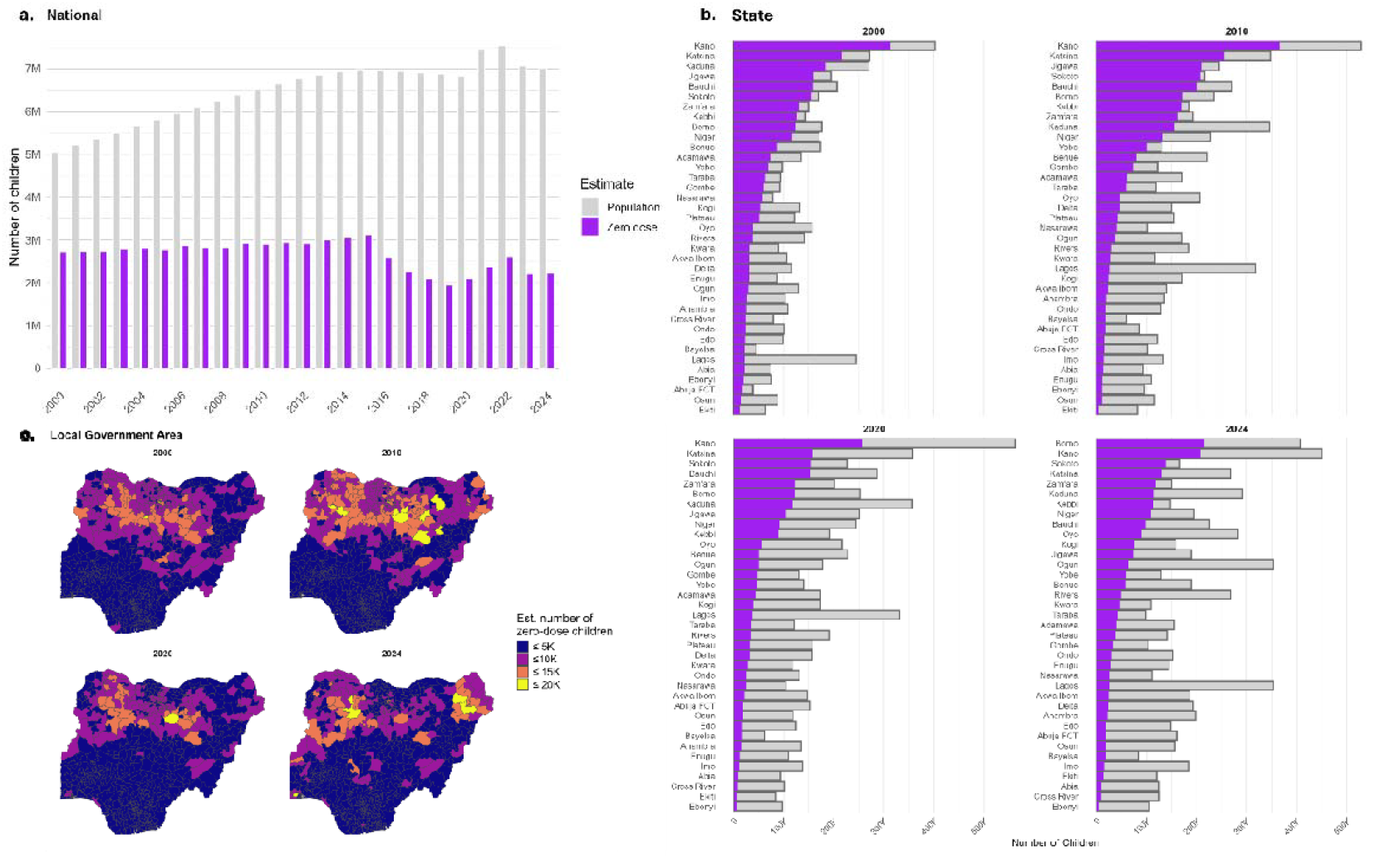
Estimates of numbers of (DTP) zero-dose children aged under 1 from 2000 to 2024 at the national, state and local government area levels.

At the state level (Figure 5b), the northeastern and northwestern states of Kano, Katsina, Kaduna, Jigawa, Sokoto, Borno, Kebbi, Zamfara and Bauchi were consistently among the states with the highest numbers of zero-dose children. Figure 5b also demonstrates the utility of the zero-dose estimates for spatial prioritization, as there can be considerable differences in the priority areas obtained using these and the population estimates. Interestingly, in 2024, Figure 5b also shows that Borno state has overtaken Kano as the state with the highest number of zero-dose children, despite the latter having a larger birth cohort.

The north-south divide in the distribution of zero-dose children is also apparent at the LGA level (Figure 5c, 6 and supplementary Figure 21), as expected, with a clear concentration of high-burden LGAs in the north. The concentration of relatively higher numbers of zero-dose children in LGAs in Borno State can also be clearly seen. However, some LGAs in the south such as those in Lagos, Ondo and Bayelsa states also had considerable numbers of zero-dose children. Figure 6 further shows that the greatest number of LGAs experiencing an increase in the number of zero-dose children was observed during 2010 – 2014, whereas the greatest reductions in zero-dose prevalence were obtained during 2015 to 2019 especially in the north (as expected). However, due to reductions in coverage post 2019, Figure 6 shows increase in numbers of zero-dose children in many LGAs both in the north and south during 2020 - 2024. Interestingly, LGAs in the northeastern state of Bauchi appear to have experienced the most reductions in zero-dose prevalence within this period. Other areas with decreases in zero-dose during this period are also concentrated in the north (e.g., Katsina, Jigawa, Gombe and Kano states) and parts of the south (e.g., Delta and Ogun states).

**Figure 6:**
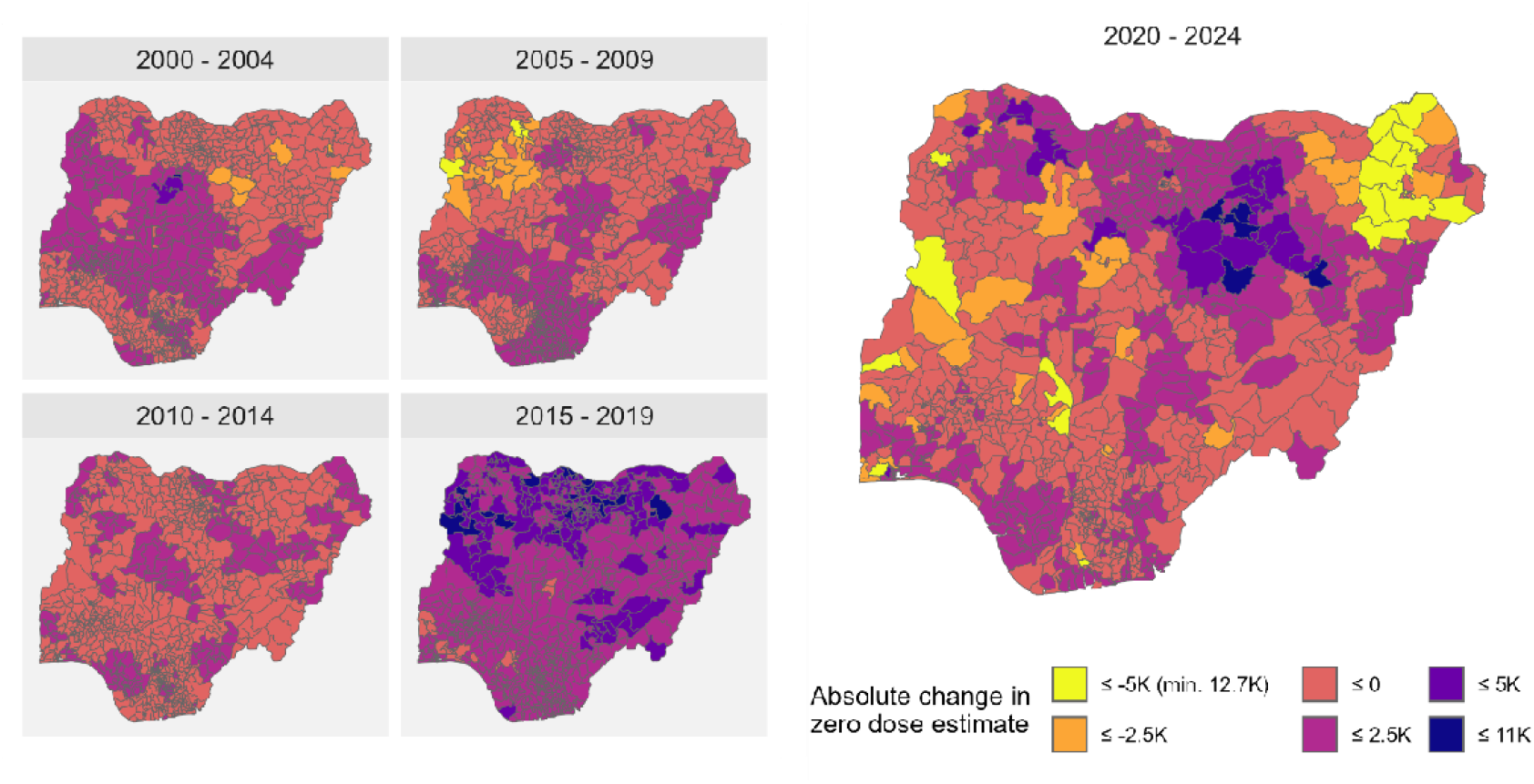
Absolute changes in estimated numbers of zero-dose children at the local government area (LGA) level from 2000 to 2014. Positive values indicate reductions in zero-dose estimates within the stated period.

## Discussion

Our study presents the most up-to-date high-resolution estimates of DTP1,3 and MCV1 coverage for Nigeria, covering an extended period from 2000 to 2024. The outputs we produced are essential for understanding programmatically important spatiotemporal dynamics in coverage. The outputs can therefore serve to guide local, targeted interventions aiming to improve coverage levels and identifying and reaching zero-dose and under-vaccinated children and missed communities within the country.

Our 1×1 km and district level estimates of coverage revealed persistent heterogeneities and a characteristic north-south divide in coverage throughout much of the study period, with a tendency for lower coverage in the north compared to the south. Between 2000 and 2015, grid-level maps showed little evidence of improvement. In contrast, noticeable gains were observed between 2015 and 2019, peaking in 2019 before declining slightly in subsequent years. These trends were similarly reflected in state and national estimates, although national averages exhibited a more gradual, long-term upward trajectory. Notably, for all three indicators, national coverage remained below 75% across the entire period, underscoring the need for intensified efforts to meet globally recognized targets of 80% for DTP1 and DTP3 and 90% for MCV1 [8].

The increases in coverage observed between 2015 and 2019 were most pronounced in the northeast and northwest, with Jigawa and Yobe demonstrating the strongest and most sustained improvements. Other historically low-coverage states in both regions - such as Kebbi, Sokoto, and Zamfara - also experienced sharp rises during this period; however, coverage in these states has since regressed to pre-2015 levels. Overall, Sokoto and Zamfara consistently exhibited the lowest coverage throughout the study period. The spatial-temporal patterns identified here are consistent with previous studies [14, 16, 18, 20, 30] and provide robust evidence to support targeted subnational strategies in Nigeria. By identifying district-level coldspots of persistently low coverage, our analyses offer a practical basis for more precise and equitable intervention planning. Moreover, successful approaches employed in states that achieved and sustained improvements (e.g., Jigawa and Yobe) could be leveraged to strengthen immunization performance in other low-coverage settings.

We found that DTP1–DTP3 dropout rates were highest in lower-coverage areas of the northeast and northwest. Although dropout rates declined in more recent years, elevated levels persisted in Sokoto, Zamfara, Borno, and Kwara - locations that also exhibited consistently low DTP1 coverage. This overlap suggests that the same structural and systemic factors driving low initial coverage also contribute to higher dropout. Consequently, areas characterized by both low DTP1 coverage and high dropout would benefit from interventions focused on improving access to vaccination services and strengthening continuity of care - particularly efforts that enhance the health system’s capacity to ensure completion of subsequent doses. Furthermore, our comparison of DTP3 and MCV1 coverage revealed relatively higher impact of vaccination campaigns on MCV1 coverage during 2015 to 2019, especially in the north. From 2021 to 2024, no noticeable effects of campaigns were observed. This may be due to the inability of DHS and MICS surveys to capture doses administered during campaigns, suboptimal campaigns or rapid loss of gains during campaigns due to underperforming RI, high birth rates, etc.

Our analysis also showed that, in the period following the COVID-19 pandemic, coverage declines were more pronounced in northern states than in the south. While previous studies have generally indicated no major national-level surge in zero-dose prevalence and have reported mixed trends at subnational levels [34, 48], our findings highlight a potential lack of resilience within northern health systems to external shocks. This underscores the need for targeted health system strengthening in these areas, a recommendation also echoed in earlier research [49]. The low coverage areas identified during this period provide a clear basis for prioritizing such focused, resilience-building efforts.

Our estimates indicate that the number of DTP zero-dose children in Nigeria consistently exceeded two million annually between 2000 and 2024, with the exception of 2019, when we estimated 1.96 million. For 2023 and 2024, we estimated 2.21 million and 2.22 million zero-dose children, respectively - figures that align closely with the 2024 WUENIC estimate of 2.1 million [3]. Importantly, unlike WUENIC, our analysis provides granular subnational insights, revealing the spatial distribution of zero-dose prevalence across states and districts. States in the northeast and northwest-particularly Kano, Katsina, Kaduna, Jigawa, Sokoto, Borno, Kebbi, Zamfara, and Bauchi-consistently harboured the highest numbers of zero-dose children. At the LGA level, the largest concentrations were also predominantly located in the north, although several LGAs in southern states such as Lagos, Ondo, and Bayelsa also exhibited substantial numbers of zero-dose children.

Our coverage and zero-dose estimates can be further integrated with complementary datasets to generate additional programmatically relevant insights. For example, linking coverage estimates with gridded data on motorized and walking travel time to the nearest health facility [23] can help quantify the number of zero-dose children living within different travel time bands. These outputs can also support assessments of coverage patterns in specific geographic contexts, including remote rural areas, urban slums, and conflict-affected settings [5]. However, such integrations must be undertaken cautiously, as the household surveys underlying our analysis may not fully capture these hard-to-reach or marginalized populations. Targeted surveys may therefore be necessary to more accurately assess coverage levels and population characteristics in these settings. For spatial prioritization, our estimates can also be combined with other programmatic indicators, such as measles and polio outbreak data - additional signals of health system weakness - to generate a more comprehensive ranking of subnational areas requiring intensified intervention.

Our study is subject to some limitations. In our methodology, DTP1 and MCV1 coverage, were more robustly estimated than DTP3 coverage. This is because the first two indicators were directly modelled, while DTP3 was derived from DTP1 and DTP3/1 to ensure internal consistency in the estimates as explained previously. Our analysis included older birth cohorts (i.e., those aged older than 12-35 months in the survey year) from surveys conducted before 2014. This has the potential to introduce recall bias in the study, with additional limitations arising from the assumption of negligible effects of catch-up vaccination, migration, and mortality, as stated previously. There is also the potential for recall bias arising from utilizing information from parental or caregiver recall in the study, although our study focused on producing crude estimates of coverage. Our estimates for Borno state should be treated with caution as the input data available for the state, particularly for the 2015, 2019 and 2020 birth cohorts were sparse due to insecurity in many areas of the state [32]. We investigated the impact of SIAs on improving MCV1 coverage through comparing combined RI and SIA MCV1 coverage with DTP3 RI coverage. However, with detailed information on the occurrence of measles SIAs, it is possible to introduce an additional indicator variable, say *z* (***s***_*i*_, *t*)in model (1), capturing whether the *t*-th birth cohort participated in an SIA or not, or a continuous variable capturing the proportion of the birth cohort that participated in an SIA. Post model-fitting, RI-specific coverage can be estimated by removing the contribution of the term from the model in both cases, see [12], for example. Lastly, our zero-dose estimates were produced using vaccination coverage and population point estimates. We will consider incorporating uncertainties from both estimates in this step in future work.

As global immunization policy increasingly prioritizes country ownership [50], governments need actionable tools and robust, timely evidence to inform strategy design and guide efficient resource allocation. In Nigeria, where substantial inequities in vaccination coverage and zero-dose prevalence persist, spatially resolved estimates offer a valuable means to target interventions more effectively. Leveraging such evidence can support more equitable access to immunization services and accelerate progress toward the goals outlined in the Immunization Agenda 2030.

## Data Availability

The data used for this work are publicly available for download from the DHS program (https://www.dhsprogram.com/), the MICS program (https://mics.unicef.org), WorldPop (www.worldpop.org) and other sources cited in this article. The authors do not have the right to redistribute these datasets. All R modelling scripts can be downloaded from https://github.com/EdsonUtazi/NGA_paper_space_time_analysis. The outputs produced by the study can be visualized here https://som-shinyapps.shinyapps.io/nga_vacn_explorer/ and can be downloaded from www.worldpop.org.

## Author contributions

CEU: Conceptualization, Data curation, Methodology, Software, Formal analysis, Visualization, Supervision, Funding acquisition, Writing – original draft, Writing – review & editing. MM: Data curation, Methodology, Software, Formal analysis, Writing – review & editing. IDO: Data curation, Visualization, Writing – review & editing. SC: Visualization, Writing – review & editing. NT-G: Conceptualization, Funding acquisition, Writing – review & editing. GM: Methodology, Writing – review & editing. YK: Methodology, Writing – review & editing. DB: Conceptualization, Resources, Writing – review & editing. JL: Conceptualization, Resources, Writing – review & editing. AJT: Conceptualization, Supervision, Funding acquisition, Writing – review & editing.

## Ethics declaration

Ethics approval for this study was obtained from the Ethics Committee at the Faculty of Environmental and Life Sciences, University of Southampton, United Kingdom (Ethics number: 50248.A1).

## References

1. World Health Organization (WHO). Immunization coverage: Key facts. 2025.

2. Shattock AJ, Johnson HC, Sim SY, Carter A, Lambach P, Hutubessy RCW, et al. Contribution of vaccination to improved survival and health: modelling 50 years of the Expanded Programme on Immunization. The Lancet. 2024;403(10441):2307–16.

3. World Health Organization. WHO/UNICEF Estimates of National Immunization Coverage (WUENIC) 2024 revision; 2024. Available from: https://www.who.int/teams/immunization-vaccines-and-biologicals/immunization-analysis-and-insights/global-monitoring/immunization-coverage/who-unicef-estimates-of-national-immunization-coverage. [Accessed on 05 July 2025].

4. Arambepola R, Yang Y, Hutchinson K, Mwansa FD, Doherty JA, Bwalya F, et al. Using geospatial models to map zero-dose children: factors associated with zero-dose vaccination status before and after a mass measles and rubella vaccination campaign in Southern province, Zambia. BMJ Global Health. 2021;6(12):e007479.

5. Wigley A, Lorin J, Hogan D, Utazi CE, Hagedorn B, Dansereau E, et al. Estimates of the number and distribution of zero-dose and under-immunised children across remote-rural, urban, and conflict-affected settings in low and middle-income countries. PLOS Global Public Health. 2022;2(10):e0001126.

6. Hogan D, Gupta A. Why Reaching Zero-Dose Children Holds the Key to Achieving the Sustainable Development Goals. Vaccines (Basel). 2023;11(4).

7. Rainey JJ, Watkins M, Ryman TK, Sandhu P, Bo A, Banerjee K. Reasons related to non-vaccination and under-vaccination of children in low and middle income countries: Findings from a systematic review of the published literature, 1999–2009. Vaccine. 2011;29(46):8215–21.

8. World Health Organization. Immunization Agenda 2030: A global strategy to leave no one behind; 2020. Available from: https://www.who.int/immunization/immunization_agenda_2030/en/. [Accessed on 25/06/2020].

9. United Nations. Transforming our world: The 2030 agenda for sustainable development; 2015. Available from: http://www.un.org/ga/search/view_doc.asp?symbol=A/RES/70/1&Lang=E. [Accessed on 20 June 2017].

10. Cutts FT, Claquin P, Danovaro-Holliday MC, Rhoda DA. Monitoring vaccination coverage: Defining the role of surveys. Vaccine. 2016;34(35):4103–9.

11. World Health Organization. World Health Organization vaccination coverage cluster surveys: reference manual; 2018. (WHO/IVB/18.09); Available from: https://apps.who.int/iris/handle/10665/272820. [Accessed on 08/12/2025].

12. Dong TQ, Wakefield J. SPACE-TIME SMOOTHING MODELS FOR SUBNATIONAL MEASLES ROUTINE IMMUNIZATION COVERAGE ESTIMATION WITH COMPLEX SURVEY DATA. Ann Appl Stat. 2021;15(4):1959–79.

13. Hancioglu A, Arnold F. Measuring Coverage in MNCH: Tracking Progress in Health for Women and Children Using DHS and MICS Household Surveys. PLOS Medicine. 2013;10(5):e1001391.

14. Utazi CE, Thorley J, Alegana VA, Ferrari MJ, Takahashi S, Metcalf CJE, et al. High resolution age-structured mapping of childhood vaccination coverage in low and middle income countries. Vaccine. 2018;36(12):1583–91.

15. Mosser JF, Gagne-Maynard W, Rao PC, Osgood-Zimmerman A, Fullman N, Graetz N, et al. Mapping diphtheria-pertussis-tetanus vaccine coverage in Africa, 2000 - 2016: a spatial and temporal modelling study. The Lancet. 2019;393(10183):1843–55.

16. Sbarra AN, Rolfe S, Nguyen JQ, Earl L, Galles NC, Marks A, et al. Mapping routine measles vaccination in low- and middle-income countries. Nature. 2021;589(7842):415–9.

17. Utazi CE, Nilsen K, Pannell O, Dotse-Gborgbortsi W, Tatem AJ. District-level estimation of vaccination coverage: Discrete vs continuous spatial models. Stat Med. 2021;40(9):2197–211.

18. Utazi CE, Thorley J, Alegana VA, Ferrari MJ, Takahashi S, Metcalf CJE, et al. Mapping vaccination coverage to explore the effects of delivery mechanisms and inform vaccination strategies. Nature Communications. 2019;10(1):1633.

19. Takahashi S, Metcalf CJE, Ferrari MJ, Tatem AJ, Lessler J. The geography of measles vaccination in the African Great Lakes region. Nature Communications. 2017;8(1):15585.

20. Dong TQ, Wakefield J. Modeling and presentation of vaccination coverage estimates using data from household surveys. Vaccine. 2021;39(18):2584–94.

21. Utazi C, Chaudhuri S, Wariri O, Olowe I, Megheib M, Tatem A. An Age-Structured Spatially Varying Coefficient Model for High Resolution Mapping of Vaccination Coverage. Preprints: Preprints; 2025.

22. Utazi CE, Yankey O, Chaudhuri S, Olowe ID, Danovaro-Holliday MC, Lazar AN, et al. Geostatistical and machine learning approaches for high-resolution mapping of vaccination coverage. Spatial and Spatio-temporal Epidemiology. 2025;54:100744.

23. Tejedor-Garavito N, Bondarenko M, Wigley A, Steingraber A, Utazi CE, Boyda D, et al. Estimating geographic access to health care services for children unvaccinated against diphtheria, tetanus, pertussis across low- and middle-income countries in 2021. medRxiv. 2025:2025.12.18.25342640.

24. Gavi Zero-Dose Learning Hub. Nigeria Zero-Dose Landscape; 2023. Available from: https://zdlh.gavi.org/. [Accessed on 16 January 2026].

25. World Health Organisation (WHO). Measles and Rubella Global Update November 2025; 2025. Available from: https://immunizationdata.who.int/global?topic=Provisional-measles-and-rubella-data&location=. [Accessed on 30 November 2025].

26. World Health Organisation (WHO). Nigeria’s Commitment to Measles and Rubella Elimination by 2030; 2025. Available from: https://www.afro.who.int/countries/nigeria/news/nigerias-commitment-measles-and-rubella-elimination-2030. [Accessed on 16 January 2026].

27. World Health Organisation (WHO). Breaking barriers, building bridges: the collaborative effort to reach every child in Nigeria; 2024. Available from: https://www.who.int/news-room/feature-stories/detail/breaking-barriers-building-bridges-collaborative-effort-nigeria. [Accessed.

28. World Health Organisation (WHO). Nigeria launches large-scale vaccination campaign to protect 106 million children against measles, rubella and polio; 2025. Available from: https://www.afro.who.int/countries/nigeria/news/nigeria-launches-large-scale-vaccination-campaign-protect-106-million-children-against-measles. [Accessed on 16 January 2026].

29. Utazi CE, Olowe ID, Chan HMT, Dotse-Gborgbortsi W, Wagai J, Umar JA, et al. Geospatial Variation in Vaccination Coverage and Zero-Dose Prevalence at the District, Ward and Health Facility Levels Before and After a Measles Vaccination Campaign in Nigeria. Vaccines (Basel). 2024;12(12).

30. Utazi CE, Wagai J, Pannell O, Cutts FT, Rhoda DA, Ferrari MJ, et al. Geospatial variation in measles vaccine coverage through routine and campaign strategies in Nigeria: Analysis of recent household surveys. Vaccine. 2020;38(14):3062–71.

31. ICF International. Demographic and Health Surveys (various) [Datasets]. Calverton, Maryland, U.S.A.: ICF International [Distributor]; 2003–2024.

32. United Nations Children’s Fund (UNICEF). Nigeria - Multiple Indicator Cluster Survey 2016/17 and 2021. 2016 – 2021.

33. World Health Organisation (WHO). Summary of measles-rubella supplementary immunization activities; 2025. Available from: https://immunizationdata.who.int/global?topic=&location=. [Accessed on 12 November 2025].

34. Aheto JMK, Olowe ID, Chan HM, Ekeh A, Dieng B, Fafunmi B, et al. Geospatial Analyses of Recent Household Surveys to Assess Changes in the Distribution of Zero-Dose Children and Their Associated Factors before and during the COVID-19 Pandemic in Nigeria. Vaccines. 2023;11(12).

35. Aheto JMK, Pannell O, Dotse-Gborgbortsi W, Trimner MK, Tatem AJ, Rhoda DA, et al. Multilevel analysis of predictors of multiple indicators of childhood vaccination in Nigeria. PLoS One. 2022;17(5):e0269066.

36. Utazi CE, Pannell O, Aheto JMK, Wigley A, Tejedor-Garavito N, Wunderlich J, et al. Assessing the characteristics of un- and under-vaccinated children in low- and middle-income countries: A multi-level cross-sectional study. PLOS Global Public Health. 2022;2(4):e0000244.

37. Tatem AJ. WorldPop, open data for spatial demography. Scientific Data. 2017;4(1):170004.

38. United Nations DoEaSA, Population Division. World Population Prospects 2024, Data Sources. UN DESA/POP/2024/DC/NO. 11; 2024. Available from: www.un.org/development/desa/pd/. [Accessed on 15 October 2025].

39. Mardia KV, Goodall CR. Spatial-temporal analysis of multivariate environmental monitoring data. Multivariate environmental statistics. 1993;6(76):347–85.

40. Moraga P. Spatial statistics for data science: theory and practice with R: Chapman and Hall/CRC; 2023.

41. Sahu S. Bayesian modeling of spatio-temporal data with R: Chapman and Hall/CRC; 2022.

42. Matérn B. Spatial Variation. 2nd ed. Berlin, Germany: Springer-Verlag; 1960.

43. Lindgren F, Rue H, Lindström J. An explicit link between Gaussian fields and Gaussian Markov random fields: the stochastic partial differential equation approach. J Roy Stat Soc Series B (Stat Methodol). 2011;73(4):423–98.

44. Utazi CE, Aheto JMK, Chan HMT, Tatem AJ, Sahu SK. Conditional probability and ratio-based approaches for mapping the coverage of multi-dose vaccines. Statistics in Medicine. 2022;41(29):5662 – 78.

45. Lindgren F, Rue H, Lindström J. Bayesian Spatial Modelling with R-INLA. Journal of Statistical Software. 2015;63(19):25.

46. Lindgren F, Bachl F, Illian J, Suen MH, Rue H, Seaton AE. inlabru: software for fitting latent Gaussian models with non-linear predictors. arXiv preprint arXiv:240700791. 2024.

47. Utazi CE, Aheto JMK, Wigley A, Tejedor-Garavito N, Bonnie A, Nnanatu CC, et al. Mapping the distribution of zero-dose children to assess the performance of vaccine delivery strategies and their relationships with measles incidence in Nigeria. Vaccine. 2023;41(1):170–81.

48. Dadari I, Sharkey A, Hoare I, Izurieta R. Analysis of the impact of COVID-19 pandemic and response on routine childhood vaccination coverage and equity in Northern Nigeria: a mixed methods study. BMJ Open. 2023;13(10):e076154.

49. Abubakar I, Dalglish SL, Angell B, Sanuade O, Abimbola S, Adamu AL, et al. The Lancet Nigeria Commission: investing in health and the future of the nation. The Lancet. 2022;399(10330):1155–200.

50. Gavi the Vaccine Alliance. A new era dawns for Gavi, as Board underlines strategic shift towards country ownership and increased support for the most vulnerable; 2025. Available from: https://bit.ly/4oxOTwk. [Accessed on 16 January 2026].

