## Supplementary for "Mapping DTP1,3 and MCV1 coverage and zero-dose prevalence in Nigeria: A spatiotemporal analysis (2000 – 2024)"

**Supplementary Information**

This supplementary file accompanies the main manuscript. It contains additional information (tables and figures) referenced in the main manuscript.

Table 1: Geolocated surveys included in the study and corresponding birth cohorts

| Survey | Dates of data collection | Age group vaccination data were obtained for | Age cohorts included in the study and corresponding birth years |
| --- | --- | --- | --- |
| 2003 DHS | March – August 2003 | 12-59 months | 12 - 23 months (2002)  24 - 35 months (2001)  36 – 48 months (2000) |
| 2008 DHS | June - October 2008 | 12-59 months | 12 - 23 months (2007)  24 - 35 months (2006)  36 – 47 months (2005)  48 – 59 months (2004) |
| 2013 DHS | February – May 2013 | 12-59 months | 12 - 23 months (2012)  24 - 35 months (2011)  36 – 47 months (2010)  48 – 59 months (2009) |
| 2016-17 MICS | September 2016-January 2017 | 12-23 months | 12 - 23 months (2015) |
| 2018 DHS | August – December 2018 | 12-35 months | 12 - 23 months (2017)  24 - 35 months (2016) |
| 2021 MICS | September – December 2021 | 12-35 months | 12 - 23 months (2020)  24 - 35 months (2019) |
| 2024 DHS | December 2023 - May 2024 | 12-35 months | 12 - 23 months (2023)  24 - 35 months (2022) |

Table 2: Measles SIA calendar for Nigeria from 2005 to 2024

| SIA type | Year | Start date | End date | Target age | Geographic coverage | Relevant birth cohorts (input data)* |
| --- | --- | --- | --- | --- | --- | --- |
| Catch-up | 2005 | 06/12/2005 | 10/12/2005 | 9 M-15 Y | National | 2000 - 2002 |
| Catch-up | 2006 | 03/10/2006 | 09/10/2006 | 9 M-15 Y | National | 2000 – 2002, 2006 |
| Follow-up | 2008 | 26/11/2008 | 15/12/2008 | 9-59 M | National | 2004 - 2007 |
| Follow-up | 2011 | 26/01/2011 | 23/02/2011 | 9-59 M | National | 2006 – 2007, 2010 |
| Follow-up | 2013 | 05/10/2013 | 09/10/2013 | 9-59 M | Subnational | 2009 - 2012 |
| Follow-up | 2013 | 02/11/2013 | 06/11/2013 | 9-59 M | Subnational | 2009 - 2012 |
| Follow-up | 2015 | 21/11/2015 | 25/11/2015 | 6 M-10 Y | Subnational | 2011, 2012, 2015 |
| Follow-up | 2016 | 28/01/2016 | 01/02/2016 | 9-59 M | National | 2015, 2012 |
| Follow-up | 2017 | 09/11/2017 | 30/04/2018 | 9-59 M | National | 2015 - 2017 |
| Follow-up | 2019 | 31/10/2019 | 30/11/2019 | 9-59 M | Subnational | 2015 – 2017, 2019 |
| Follow-up | 2020 | 09/10/2020 | 18/10/2020 | 9-59 M | Subnational | 2016, 2017, 2019, 2020 |
| Follow-up | 2021 | 01/11/2021 | 10/11/2021 | 9-59 M | Subnational | 2017, 2019, 2020 |
| Follow-up | 2022 | 01/06/2022 | 10/06/2022 | 9-59 M | Subnational | 2017, 2019-2020 |
| Follow-up | 2023 | 2023-10 | 2023-12 | 9-59 M | Subnational | 2019-2020, 2022 - 2023 |
| Follow-up | 2024 | 15/07/2024 | 27/07/2024 | 9-59 M | Subnational | 2020, 2022-2023 |

*****The determination of the birth cohorts in the input data covered by these SIAs is approximate and not exact.

Table 3: DHS variables included in the study and detailed definitions

| Covariate name*  (short name) | Description | Sources | Interpolation method |
| --- | --- | --- | --- |
| Household wealth  (Wealth) | Proportion of households classified as middle/richer/richest according to DHS classification | 2003, 2008, 2013, 2018 and 2024 Nigeria Demographic and Health Survey | See methods section |
| Maternal education  (Education) | Proportion of mothers who had at least a primary education | 2003, 2008, 2013, 2018 and 2024 Nigeria Demographic and Health Survey | See methods section |
| Skilled birth attendance  (SBA) | Proportion of live births in the 2 years preceding the survey that were assisted by a skilled provider (doctor, nurse/midwife or auxiliary midwife) | 2003, 2008, 2013, 2018 and 2024 Nigeria Demographic and Health Survey | See methods section |
| Health card/document ownership  (Health card) | Proportion of children who owned a vaccination card or document (either seen or unseen) during the survey | 2003, 2008, 2013, 2018 and 2024 Nigeria Demographic and Health Survey | See methods section |
| Proportion not stunted  (Not stunted) | Compliment of proportion of eligible children age 0-59 months with height-for-age z-score < -2 SD below the WHO median for their age and sex. | 2003, 2008, 2013, 2018 and 2024 Nigeria Demographic and Health Survey | See methods section |
| Access to media  (Media) | Proportion of households with access to newspaper, radio or television. | 2003, 2008, 2013, 2018 and 2024 Nigeria Demographic and Health Survey | See methods section |

*These covariates were calculated at the cluster level in our analyses by joining the household and women’s file to the children’s file and then subsetting the resulting data to children who were alive during each survey.

Table 4: Geospatial covariates included in the study

| Covariate description | Temporal coverage | Spatial resolution | Reference |
| --- | --- | --- | --- |
| Enhanced vegetation index  (EVI) | 2000-2024 | 1km | Didan, K. (2021). MODIS/Terra Vegetation Indices Monthly L3 Global 1km SIN Grid V061 [Data set]. NASA EOSDIS Land Processes Distributed Active Archive Center. Accessed 2025-03-03 from https://doi.org/10.5067/MODIS/MOD13A3.061 |
| Parasite rate of plasmodium falciparum (PfPR) in children between the ages of 2 and 10 years old  (Malaria prev.) | 2000-2022 | 1km | Weiss DJ, Lucas TCD, Nguyen M, et al. Mapping the global prevalence, incidence, and mortality of Plasmodium falciparum, 2000–17: a spatial and temporal modelling study. Lancet 2019; published online June 19. DOI: 10.1016/S0140-6736(19)31097-9 Battle KE, Lucas TCD, Nguyen M, et al. Mapping the global endemicity and clinical burden of Plasmodium vivax, 2000–17: a spatial and temporal modelling study. Lancet 2019; published online June 19. DOI: 10.1016/S0140-6736(19)31096-7 https://malariaatlas.org/explorer/#/ |
| Annual average Maximum temperature (Degree Celsius)  (Max. temp) | 2000-2024 | 1km | Abatzoglou, J.T., S.Z. Dobrowski, S.A. Parks, K.C. Hegewisch, 2018, Terraclimate, a high-resolution global dataset of monthly climate and climatic water balance from 1958-2015, Scientific Data |
| Annual average precipitation (mm/year)  (Precipitation) | 2000-2024 | 1km | Abatzoglou, J.T., S.Z. Dobrowski, S.A. Parks, K.C. Hegewisch, 2018, Terraclimate, a high-resolution global dataset of monthly climate and climatic water balance from 1958-2015, Scientific Data |
| Distance to Urban Areas (km)  (Dist. to urban areas) | 2000-2022 | 1km | Copernicus Climate Change Service, Climate Data Store, (2019): Land cover classification gridded maps from 1992 to present derived from satellite observation. Copernicus Climate Change Service (C3S) Climate Data Store (CDS). DOI: 10.24381/cds.006f2c9a |

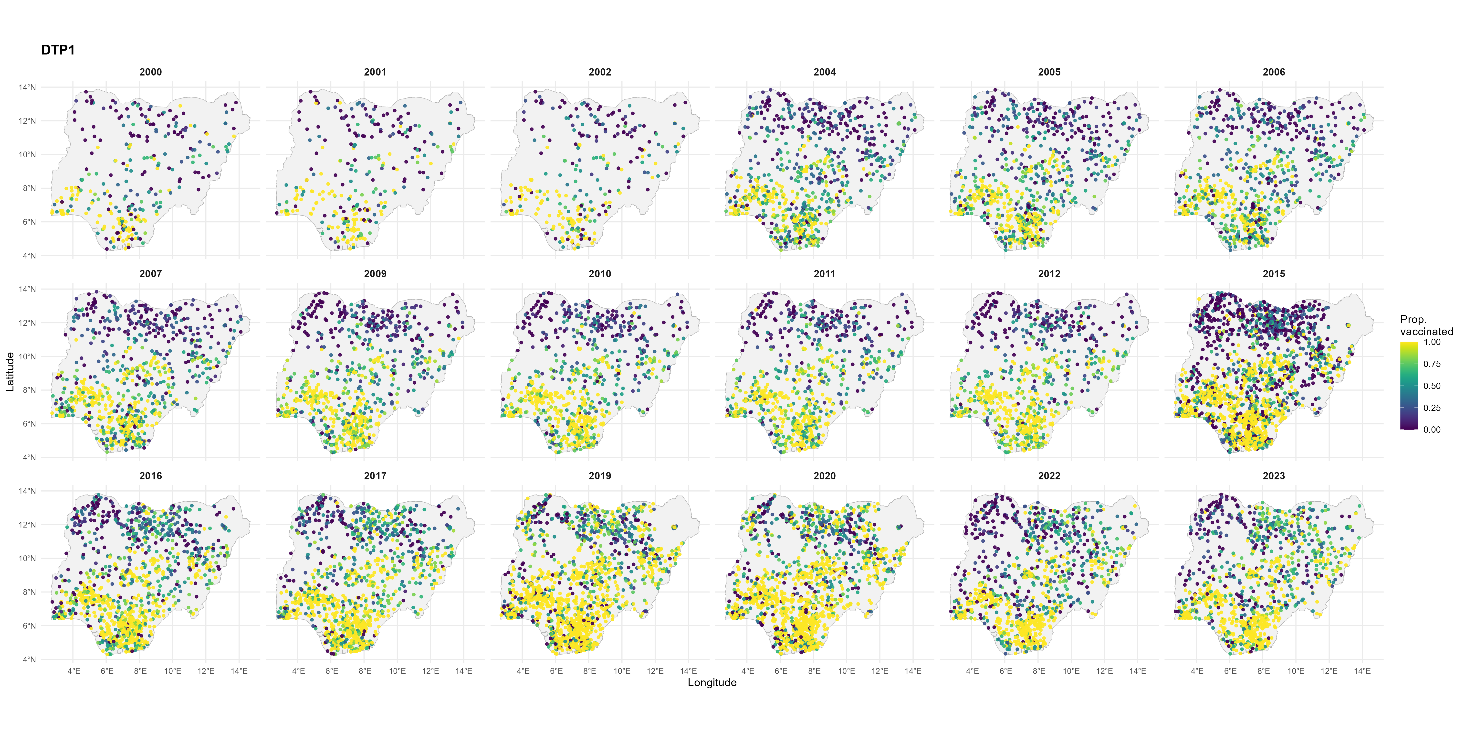

Figure 1: Cluster-level data for DTP1. The years shown correspond to the birth years of the age cohorts included in the study (see Supplementary Table 1).

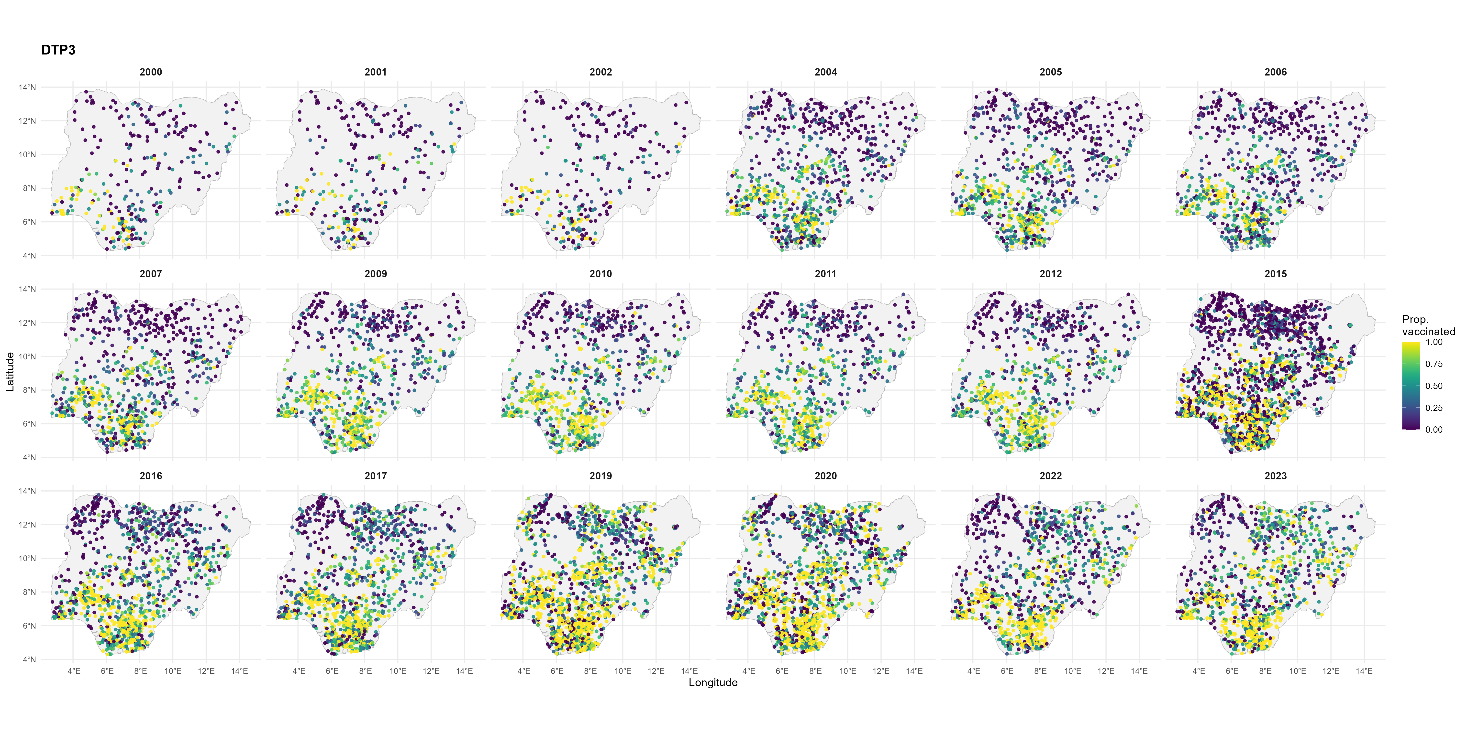

Figure 2: Cluster-level data for DTP3. The years shown correspond to the birth years of the age cohorts included in the study (see Supplementary Table 1).

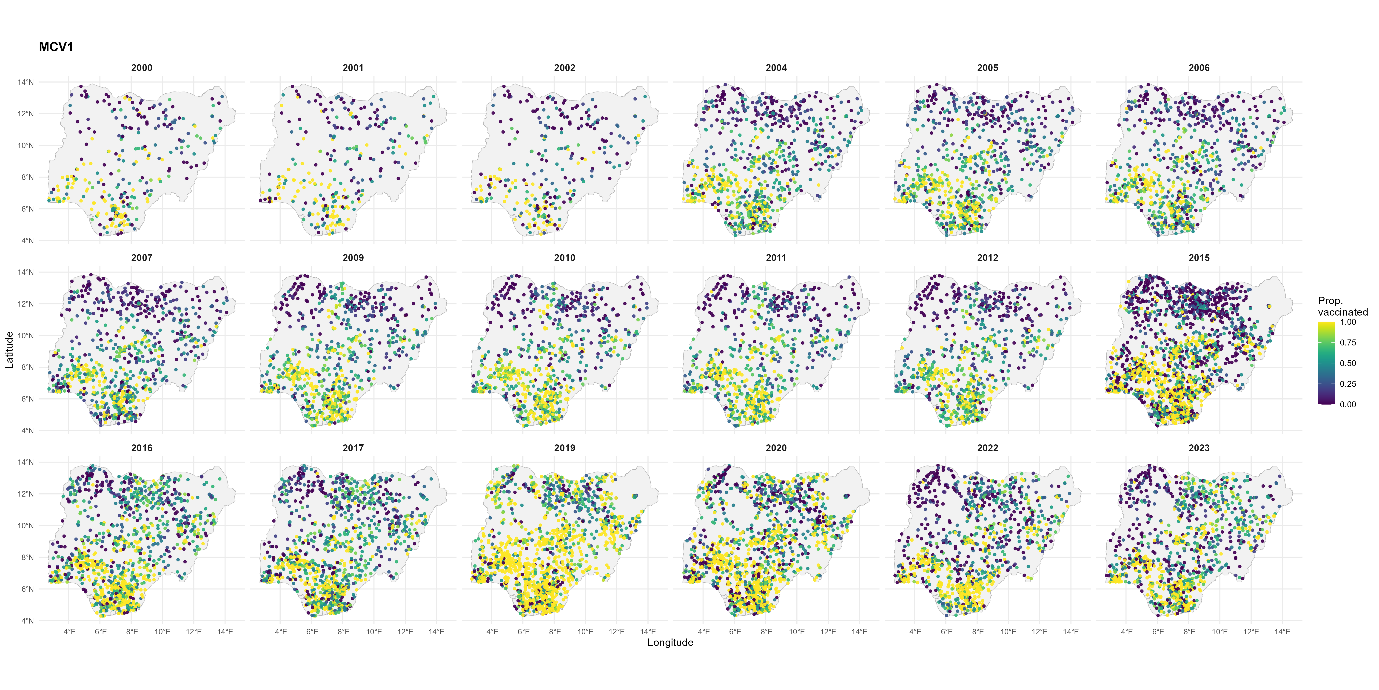

Figure 3: Cluster-level data for MCV1. The years shown correspond to the birth years of the age cohorts included in the study (see Supplementary Table 1).

Table 5: Parameter estimates for DTP1

| **Parameter** | **Mean** | **Std. dev.** | **2.5%** | **97.5%** |
| --- | --- | --- | --- | --- |
| Intercept | -0.691 | 0.682 | -2.027 | 0.645 |
| Wealth | 0.048 | 0.104 | -0.157 | 0.252 |
| Education | 0.916 | 0.139 | 0.644 | 1.188 |
| SBA | 0.749 | 0.152 | 0.451 | 1.047 |
| Health card | 4.653 | 0.145 | 4.369 | 4.937 |
| Not stunted | -0.365 | 0.229 | -0.814 | 0.083 |
| Media | 0.158 | 0.133 | -0.103 | 0.42 |
| Dist. to urban areas | -0.004 | 0.002 | -0.008 | -0.001 |
| EVI | -1.400 | 0.33 | -2.048 | -0.753 |
| Max. temperature | -0.023 | 0.019 | -0.061 | 0.016 |
| Precipitation | 0.143 | 0.111 | -0.075 | 0.36 |
| Malaria prevalence | -1.432 | 0.267 | -1.955 | -0.909 |
| Spatial range for $\omega$ $(r)$* | 2.06 | 0.194 | 1.713 | 2.475 |
| Std. dev. for $\omega$ $(\sigma_{s})$ | 1.017 | 0.058 | 0.91 | 1.138 |
| Autocorrelation for $\omega$ $(\rho)$ | 0.857 | 0.02 | 0.816 | 0.894 |
| Precision for $\epsilon(\sigma_{\epsilon}^{-2})$ | 2.036 | 0.113 | 1.822 | 2.267 |

*Estimated range is 228 km

Table 6: Parameter estimates for DTP3/1

| **Parameter** | **Mean** | **Std. dev.** | **2.5%** | **97.5%** |
| --- | --- | --- | --- | --- |
| Intercept | -3.894 | 1.102 | -6.054 | -1.734 |
| Wealth | 0.796 | 0.179 | 0.445 | 1.146 |
| Education | 0.31 | 0.24 | -0.161 | 0.78 |
| SBA | 0.722 | 0.241 | 0.249 | 1.195 |
| Health card | 2.927 | 0.236 | 2.463 | 3.39 |
| Not stunted | 0.315 | 0.356 | -0.384 | 1.013 |
| Media | -0.216 | 0.216 | -0.639 | 0.207 |
| Dist. to urban areas | 0.0005 | 0.003 | -0.006 | 0.005 |
| EVI | -0.515 | 0.506 | -1.506 | 0.477 |
| Max. temperature | 0.102 | 0.032 | 0.039 | 0.165 |
| Precipitation | -0.007 | 0.169 | -0.337 | 0.323 |
| Malaria prevalence | -1.401 | 0.454 | -2.292 | -0.51 |
| Spatial range for $\omega$ $(r)$* | 1.648 | 0.16 | 1.357 | 1.988 |
| Std. dev. for $\omega$ $(\sigma_{s})$ | 1.505 | 0.07 | 1.373 | 1.648 |
| Autocorrelation for $\omega$ $(\rho)$ | 0.775 | 0.025 | 0.722 | 0.822 |
| Precision for $\epsilon(\sigma_{\epsilon}^{-2})$ | 0.321 | 0.011 | 0.3 | 0.344 |

*Estimated range is 183 km

Table 7: Parameter estimates for MCV1

| **Parameter** | **Mean** | **Std. dev.** | **2.5%** | **97.5%** |
| --- | --- | --- | --- | --- |
| Intercept | -2.216 | 0.785 | -3.754 | -0.678 |
| Wealth | 0.105 | 0.095 | -0.081 | 0.29 |
| Education | 0.819 | 0.127 | 0.57 | 1.069 |
| SBA | 0.875 | 0.133 | 0.614 | 1.136 |
| Health card | 3.218 | 0.128 | 2.966 | 3.469 |
| Not stunted | 0.33 | 0.203 | -0.068 | 0.728 |
| Media | 0.243 | 0.117 | 0.014 | 0.472 |
| Dist. to urban areas | -0.003 | 0.002 | -0.006 | 0.0004 |
| EVI | -1.315 | 0.292 | -1.887 | -0.743 |
| Max. temperature | 0.017 | 0.022 | -0.027 | 0.06 |
| Precipitation | 0.152 | 0.125 | -0.093 | 0.398 |
| Malaria prevalence | -1.179 | 0.262 | -1.692 | -0.665 |
| Spatial range for $\omega$ $(r)$* | 2.592 | 0.198 | 2.228 | 3.007 |
| Std. dev. for $\omega$ $(\sigma_{s})$ | 1.129 | 0.066 | 1.006 | 1.267 |
| Autocorrelation for $\omega$ $(\rho)$ | 0.832 | 0.021 | 0.788 | 0.871 |
| Precision for $\epsilon(\sigma_{\epsilon}^{-2})$ | 2.342 | 0.127 | 2.104 | 2.603 |

*Estimated range is 287 km

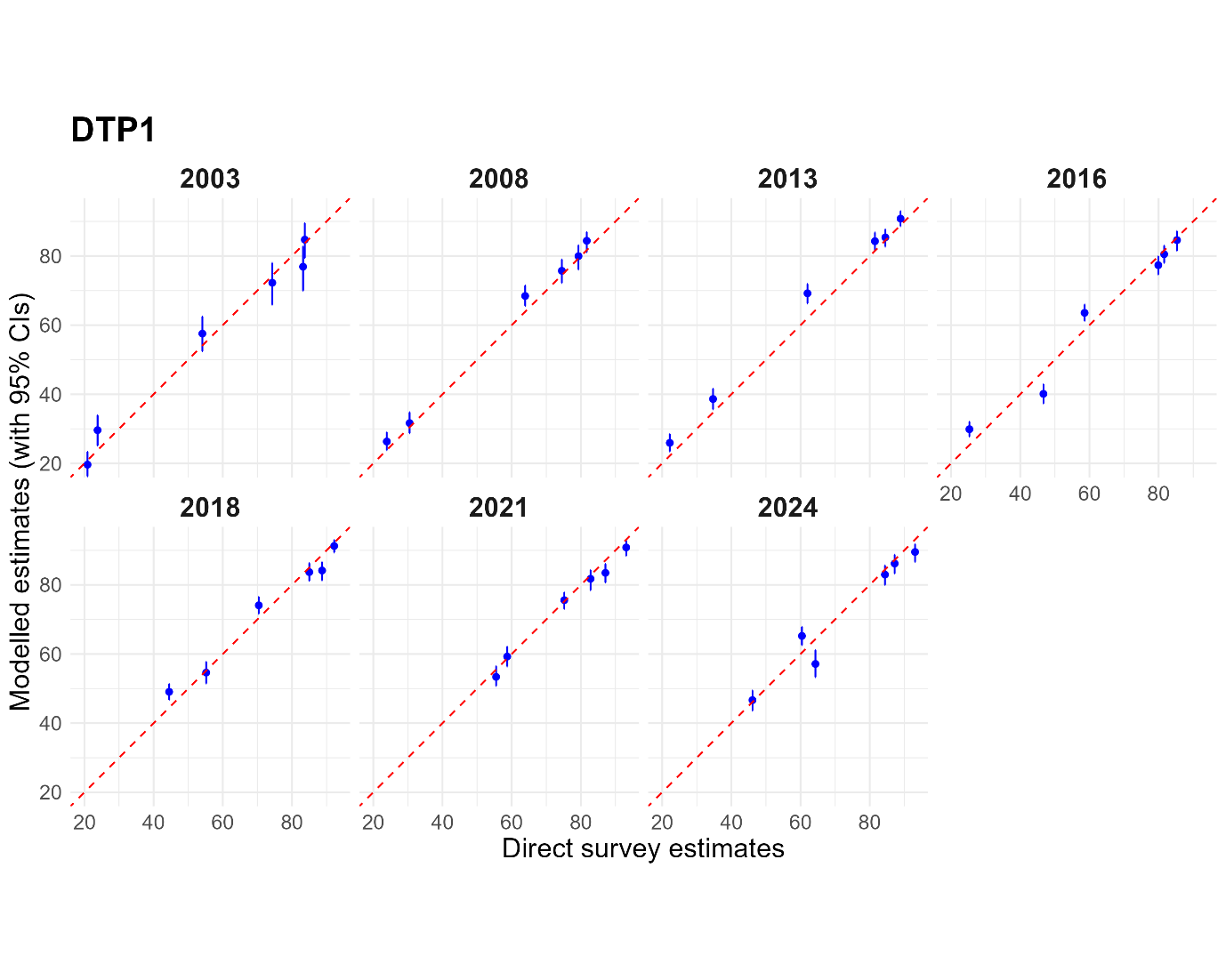

Figure 4: Plots of direct survey estimates versus modelled estimates at the regional level for DTP1 across the survey years. The direct survey estimates shown are for children aged 12 – 23 months in the survey year. These are matched up with the modelled estimates produced for the year preceding each survey year, corresponding to the same birth cohort.

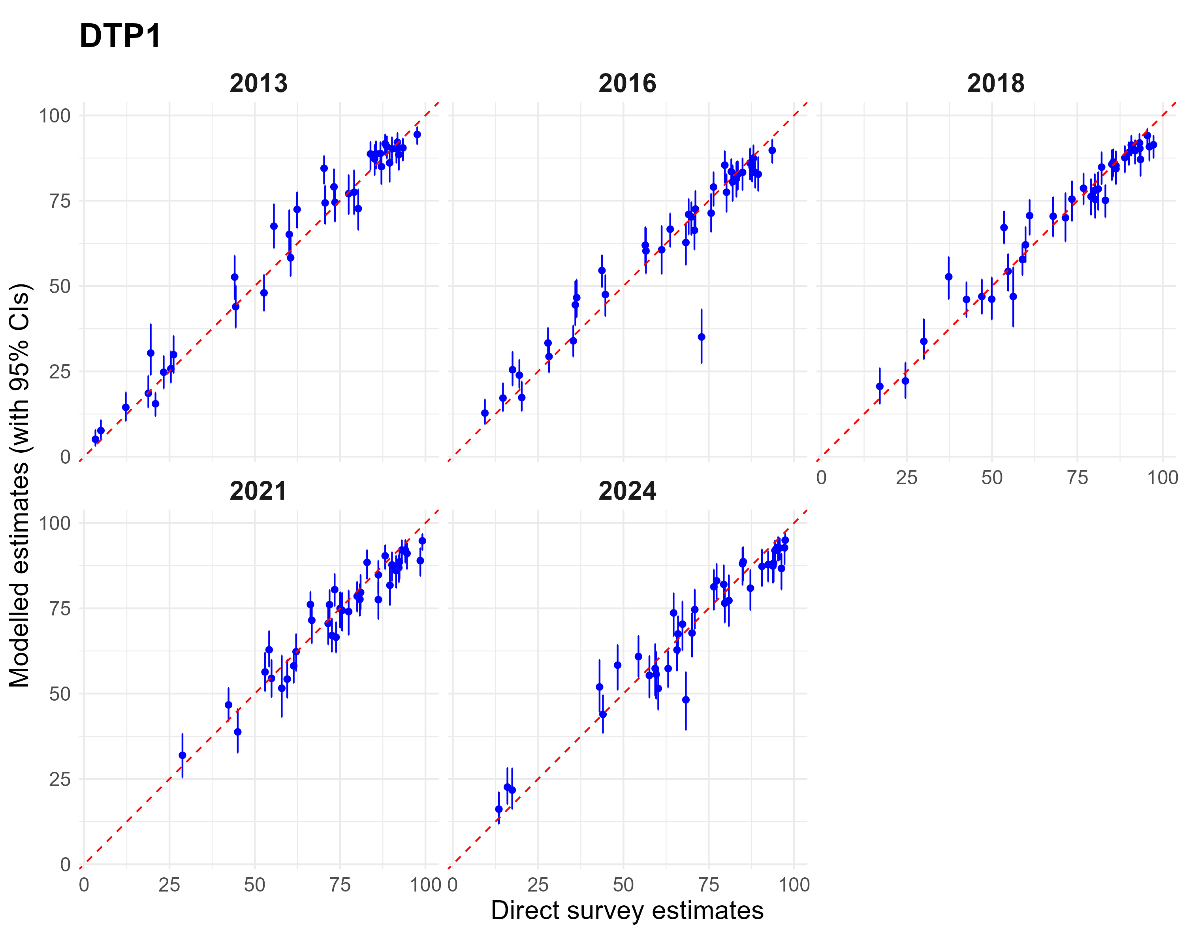

Figure 5: Plots of direct survey estimates versus modelled estimates at the state level for DTP1 across the survey years. The direct survey estimates shown are for children aged 12 – 23 months in the survey year. These are matched up with the modelled estimates produced for the year preceding each survey year, corresponding to the same birth cohort.

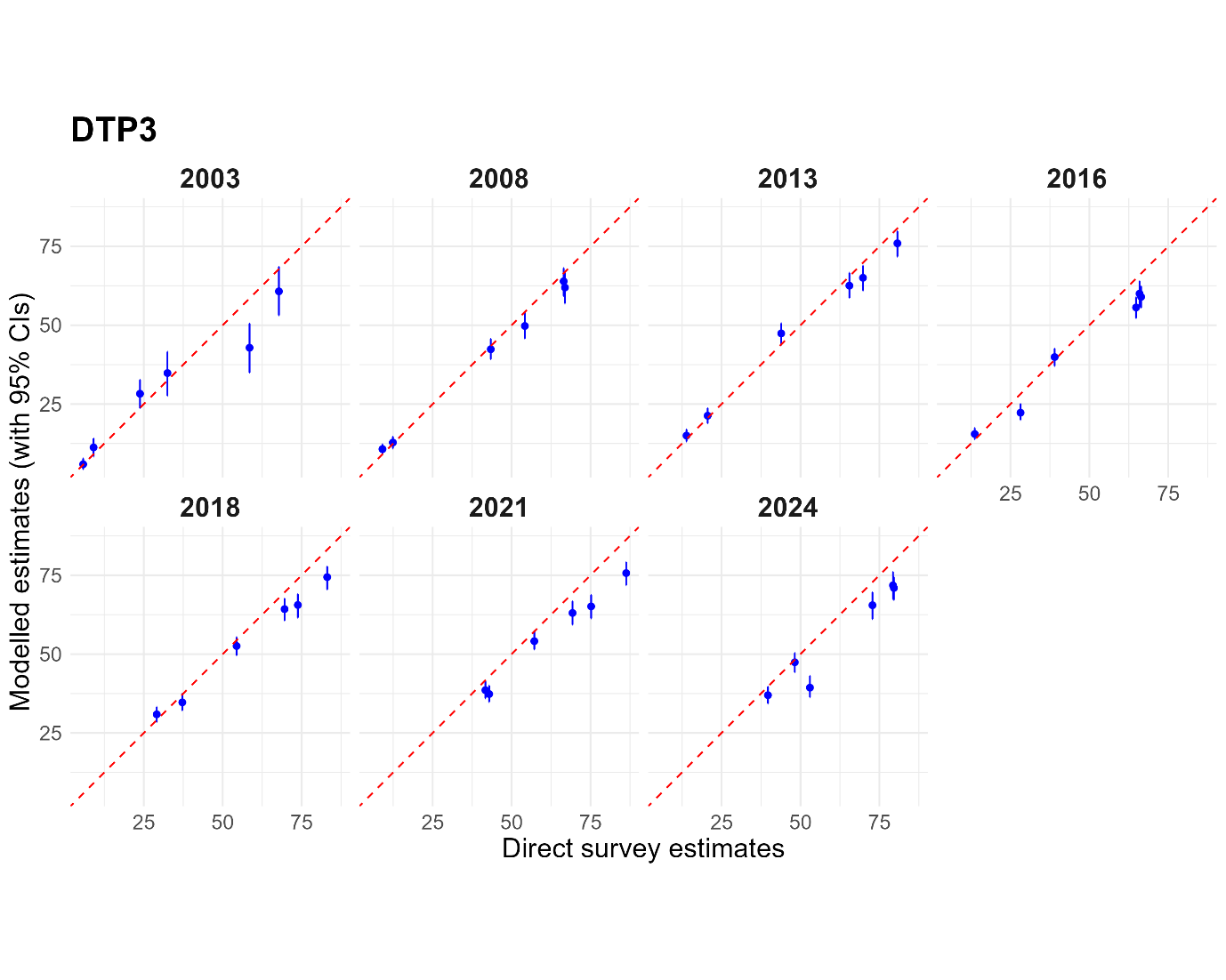

Figure 6: Plots of direct survey estimates versus modelled estimates at the regional level for DTP3 across the survey years. The direct survey estimates shown are for children aged 12 – 23 months in the survey year. These are matched up with the modelled estimates produced for the year preceding each survey year, corresponding to the same birth cohort.

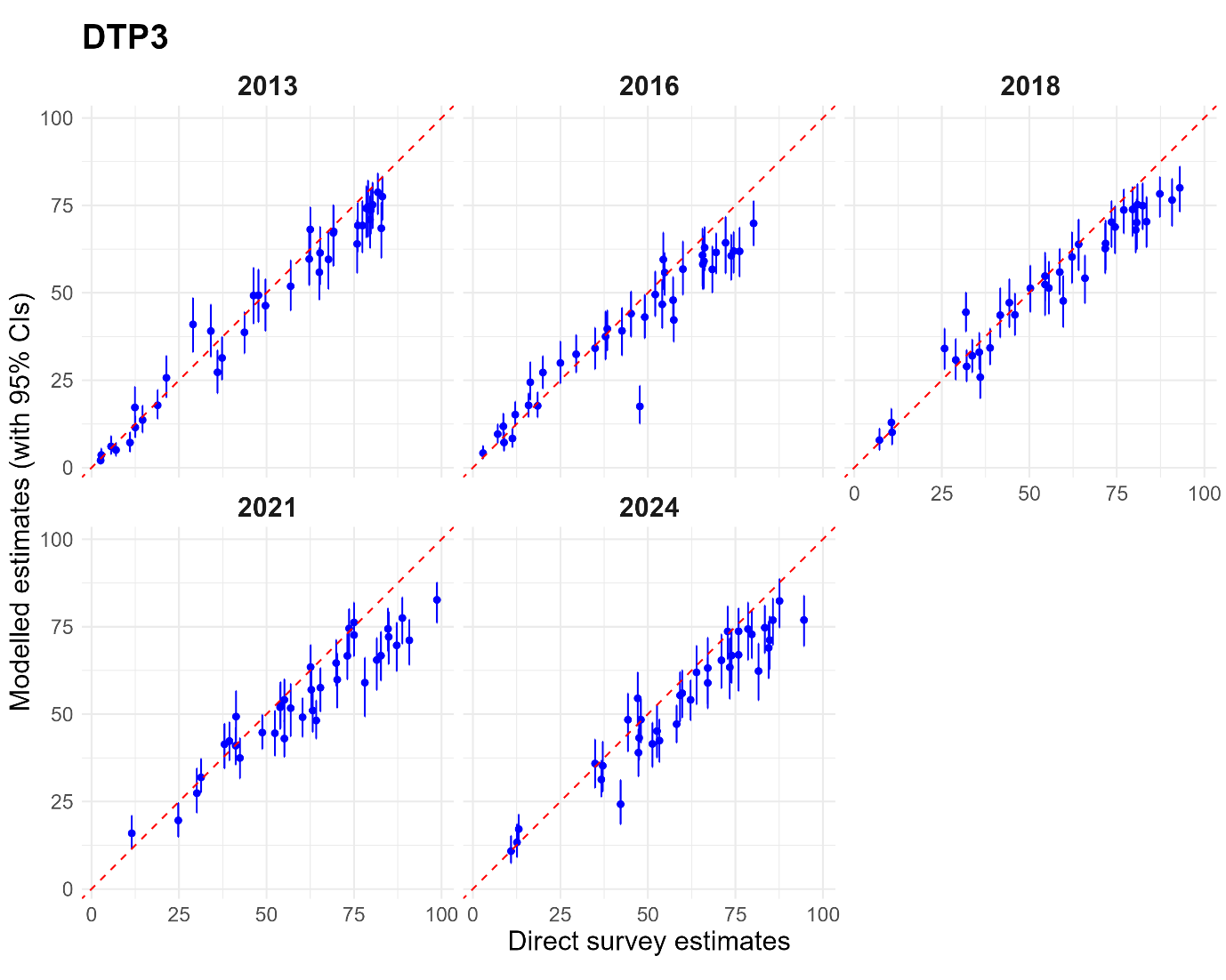

Figure 7: Plots of direct survey estimates versus modelled estimates at the state level for DTP3 across the survey years. The direct survey estimates shown are for children aged 12 – 23 months in the survey year. These are matched up with the modelled estimates produced for the year preceding each survey year, corresponding to the same birth cohort.

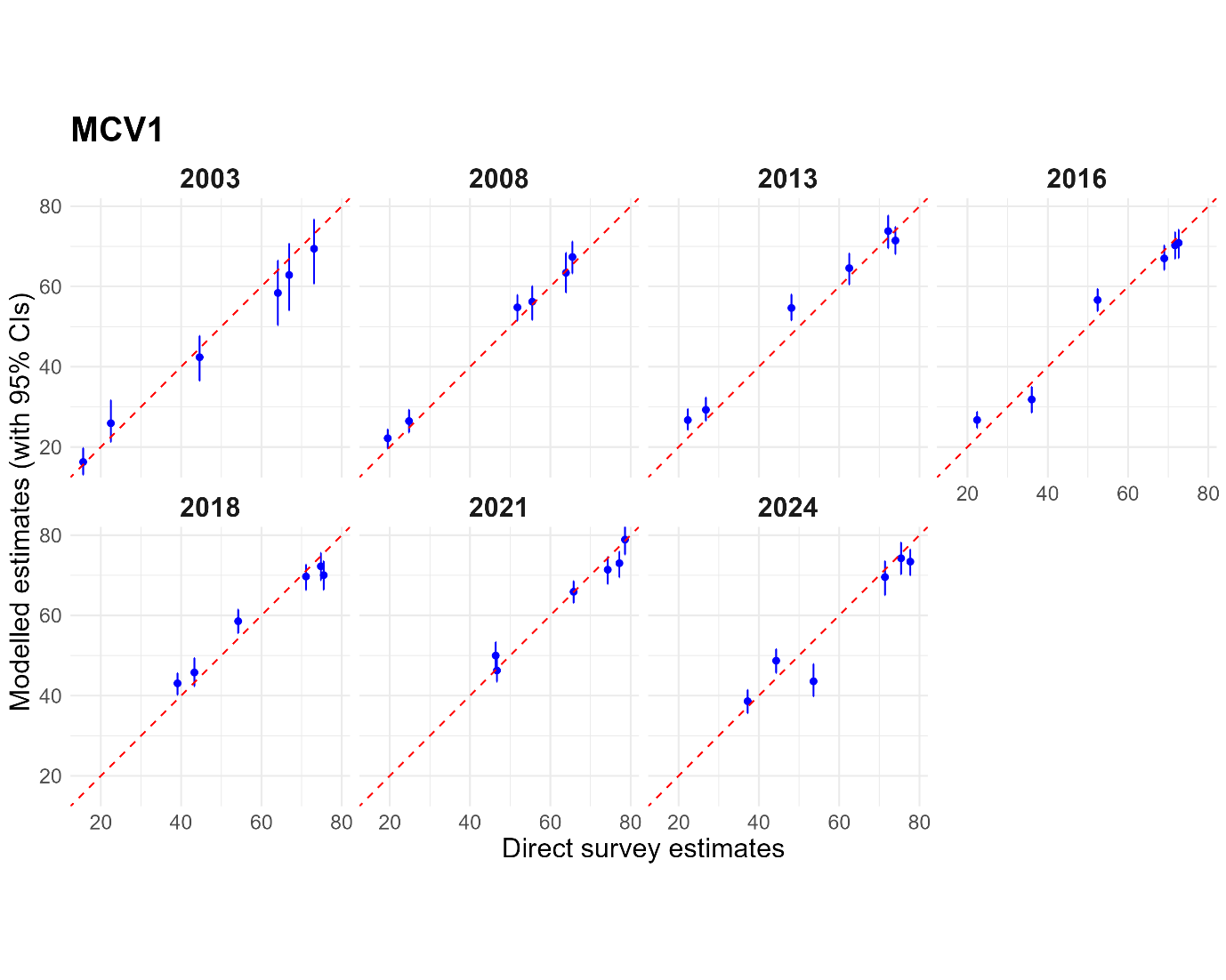

Figure 8: Plots of direct survey estimates versus modelled estimates at the regional level for MCV1 across the survey years. The direct survey estimates shown are for children aged 12 – 23 months in the survey year. These are matched up with the modelled estimates produced for the year preceding each survey year, corresponding to the same birth cohort.

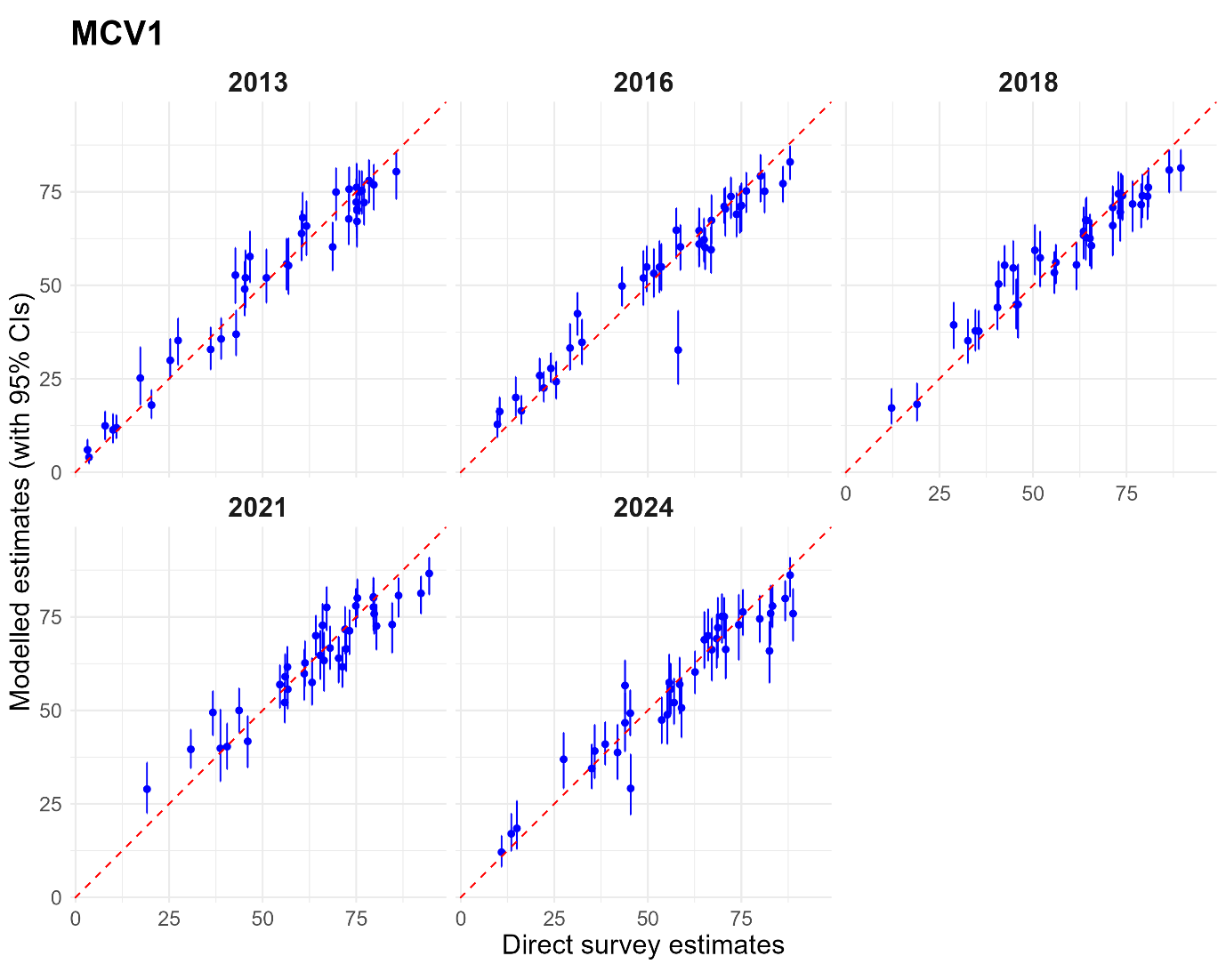

Figure 9: Plots of direct survey estimates versus modelled estimates at the regional level for DTP1 across the survey years. The direct survey estimates shown are for children aged 12 – 23 months in the survey year. These are matched up with the modelled estimates produced for the year preceding each survey year, corresponding to the same birth cohort.

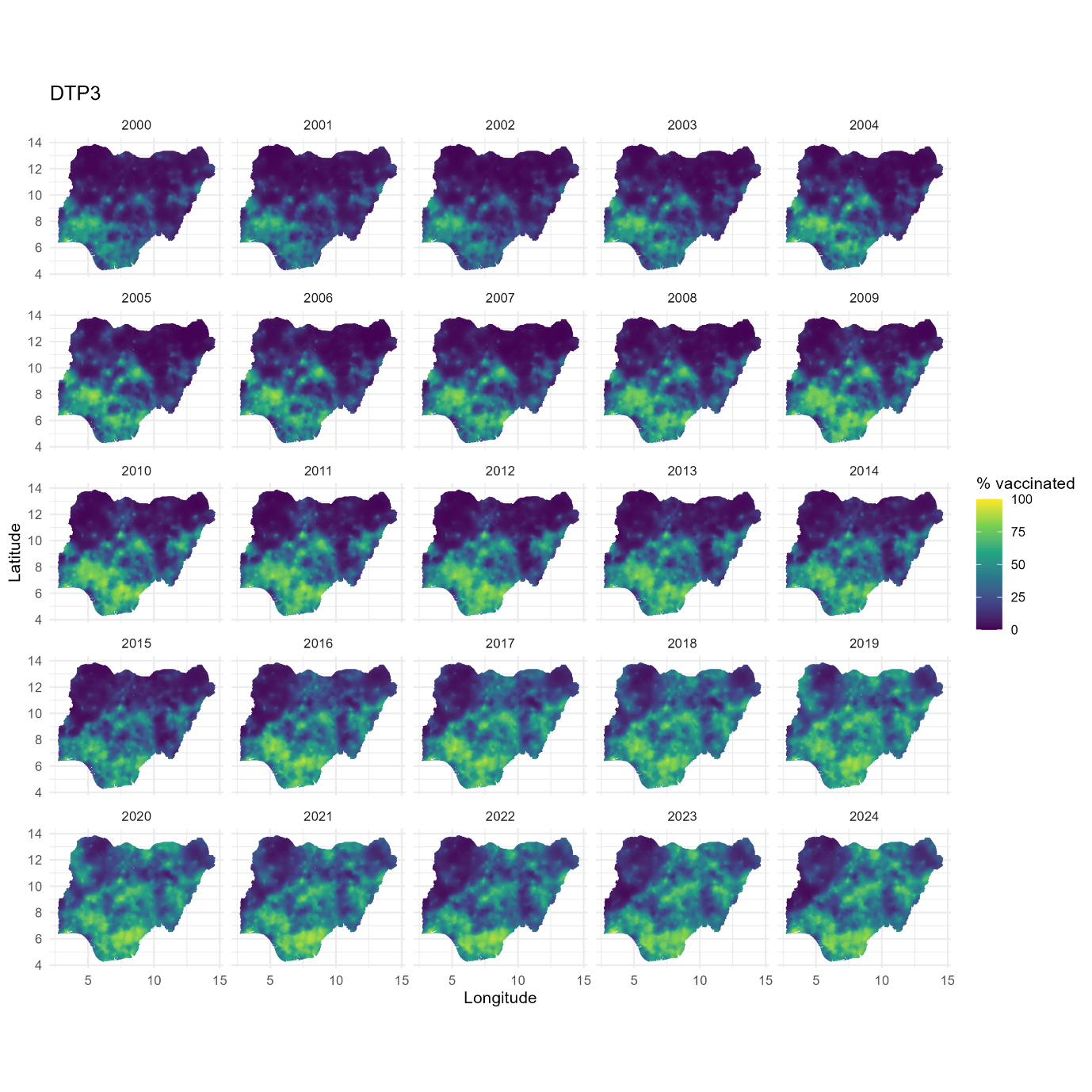

Figure 10: 1x1 km estimates of DTP3 coverage for Nigeria from 2000 to 2024. These estimates relate to the birth cohort in each year. The associated uncertainties are shown in supplementary Figure 13.

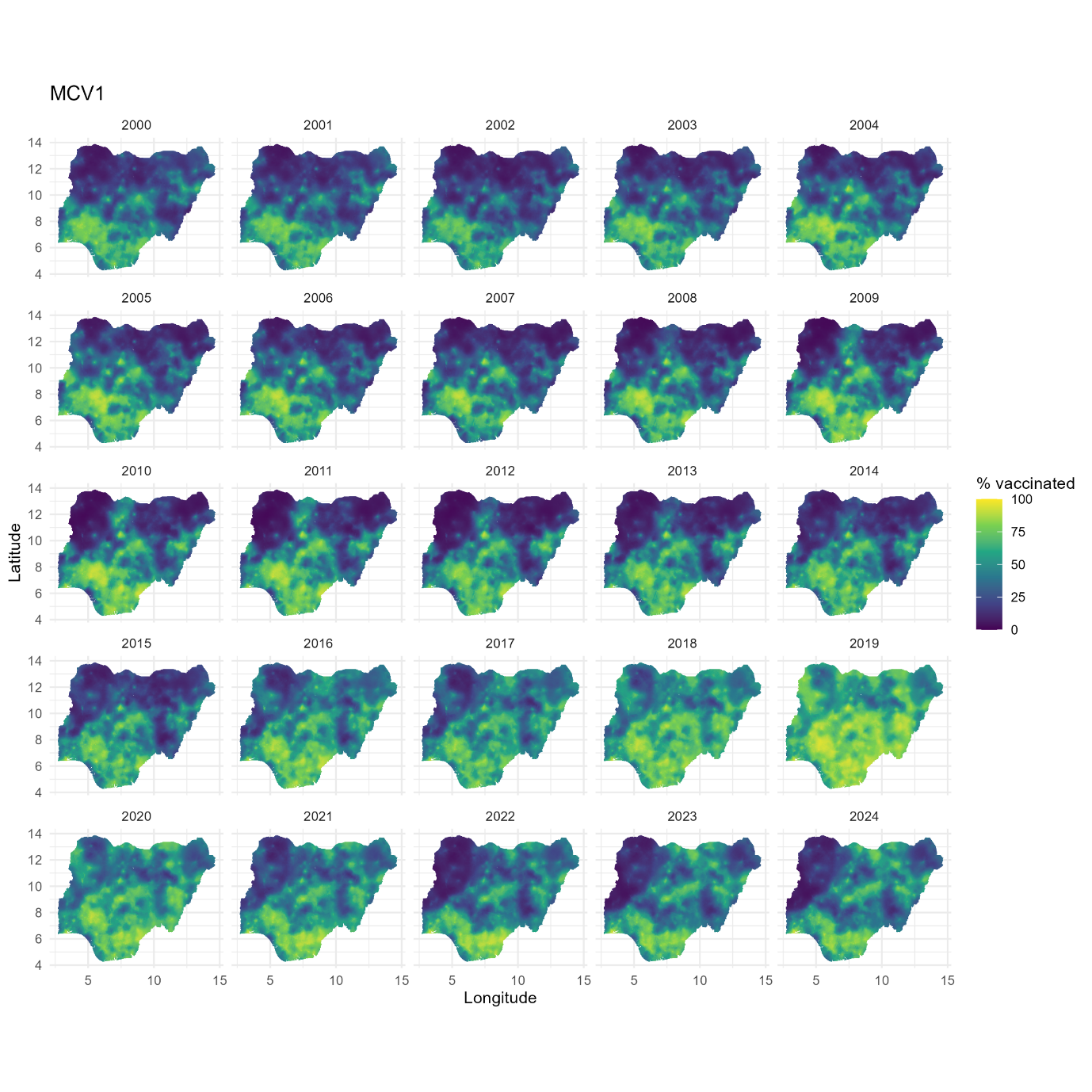

Figure 11: 1x1 km estimates of MCV1 coverage for Nigeria from 2000 to 2024. These estimates relate to the birth cohort in each year. The associated uncertainties are displayed in supplementary Figure 14.

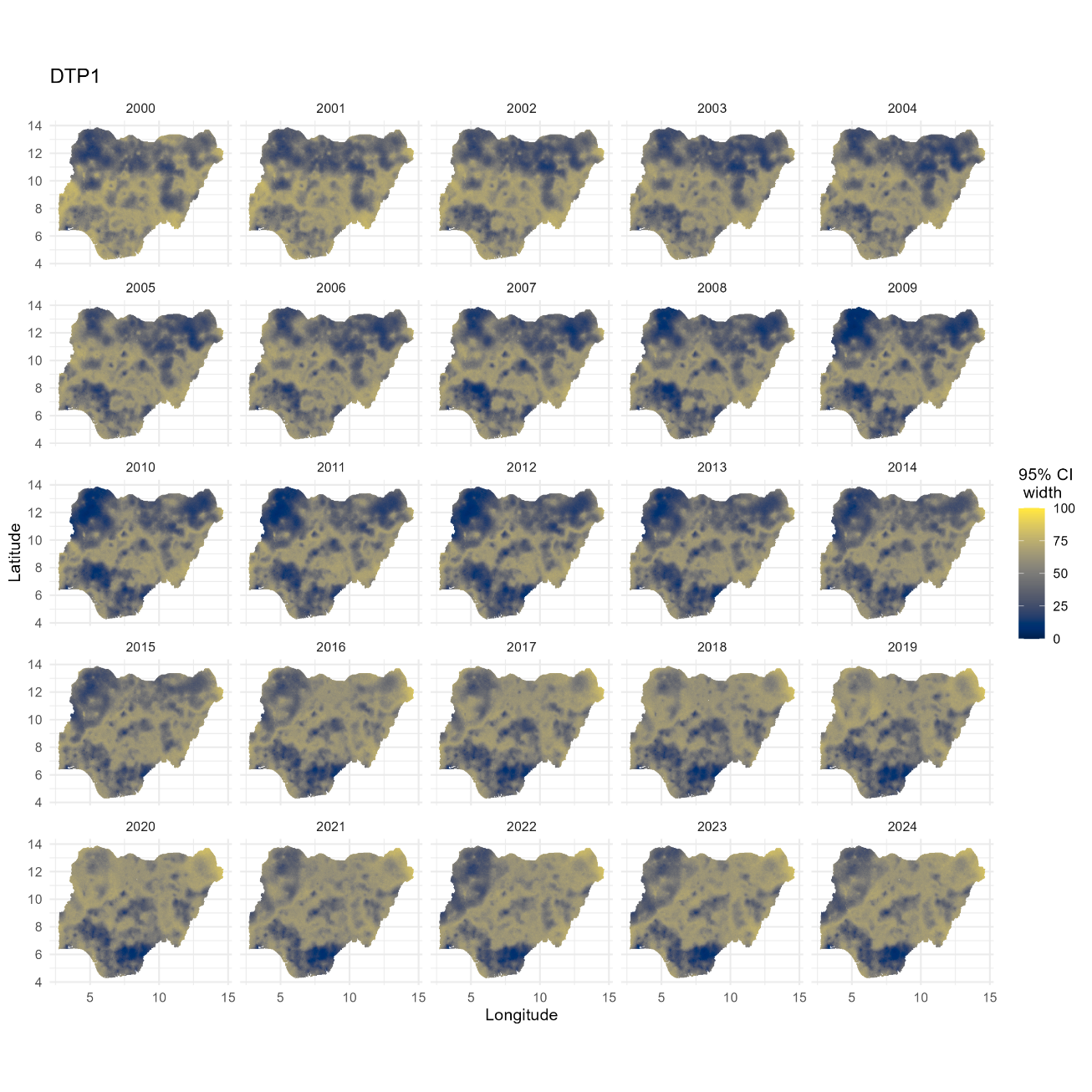

Figure 12: 1x1 km uncertainty estimates for DTP1 from 2000 to 2024, shown as the widths of the 95% credible intervals. These estimates relate to the birth cohort in each year.

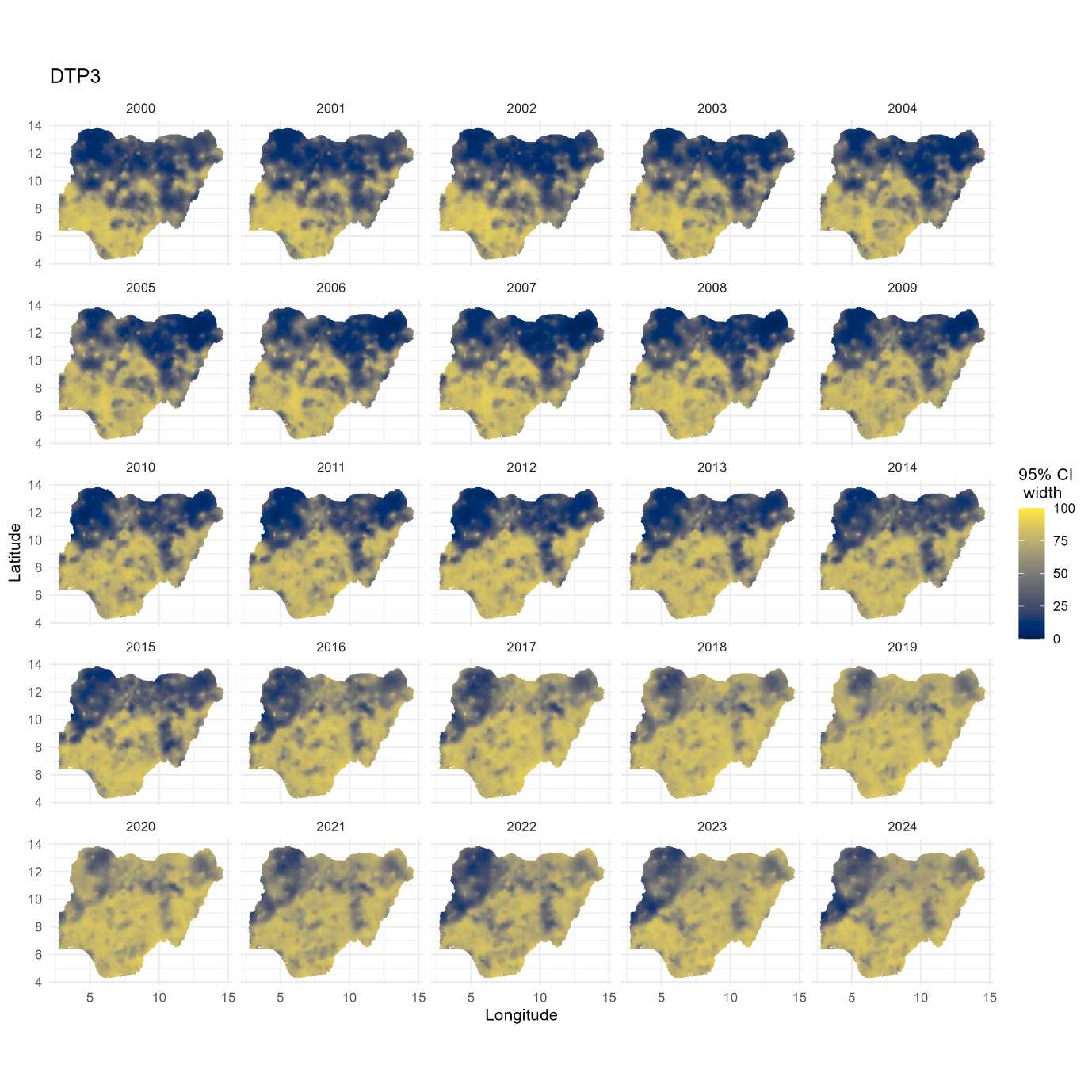

Figure 13: 1x1 km uncertainty estimates for DTP3 from 2000 to 2024, shown as the widths of the 95% credible intervals. These estimates relate to the birth cohort in each year.

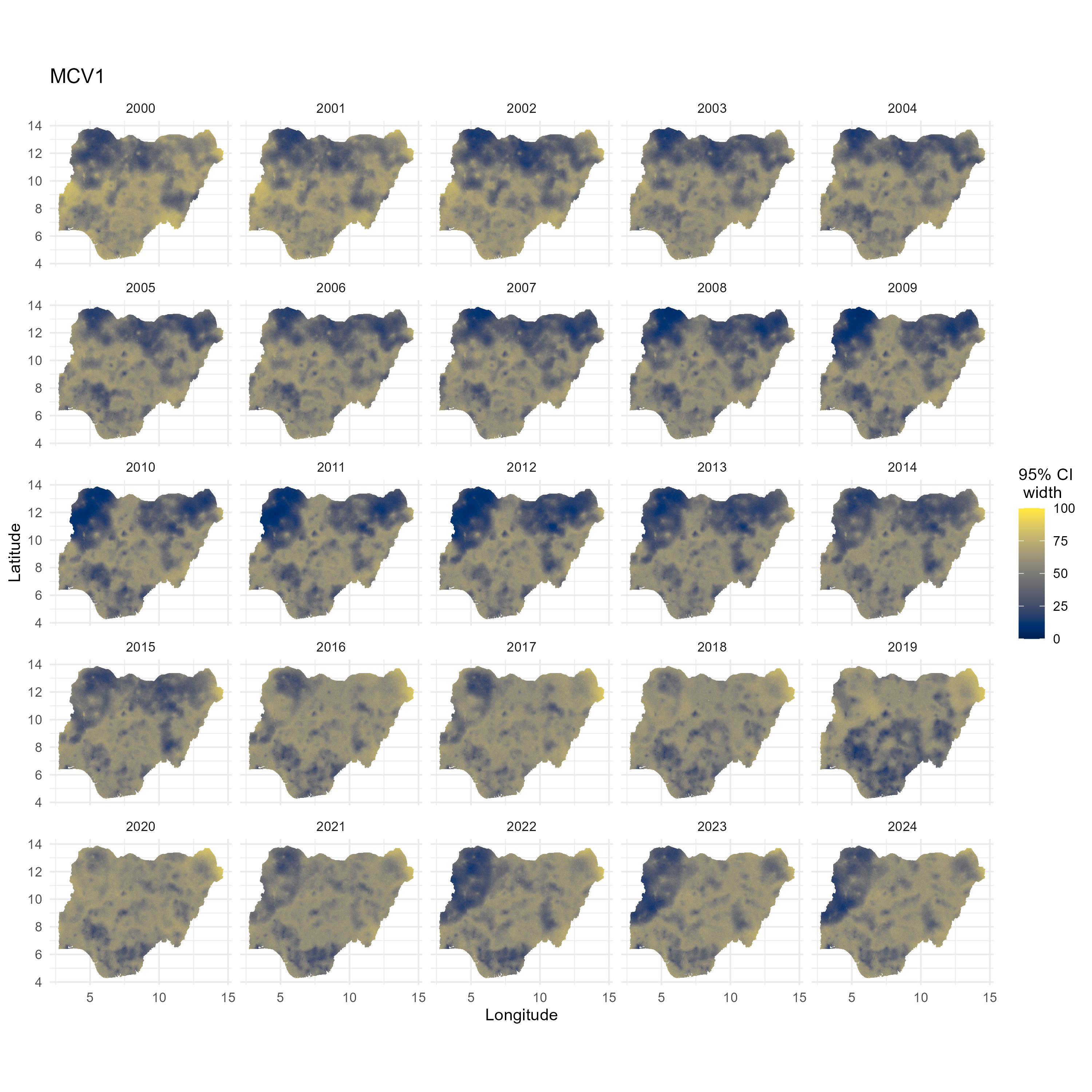

Figure 14: 1x1 km uncertainty estimates for MCV1 from 2000 to 2024, shown as the widths of the 95% credible intervals. These estimates relate to the birth cohort in each year.

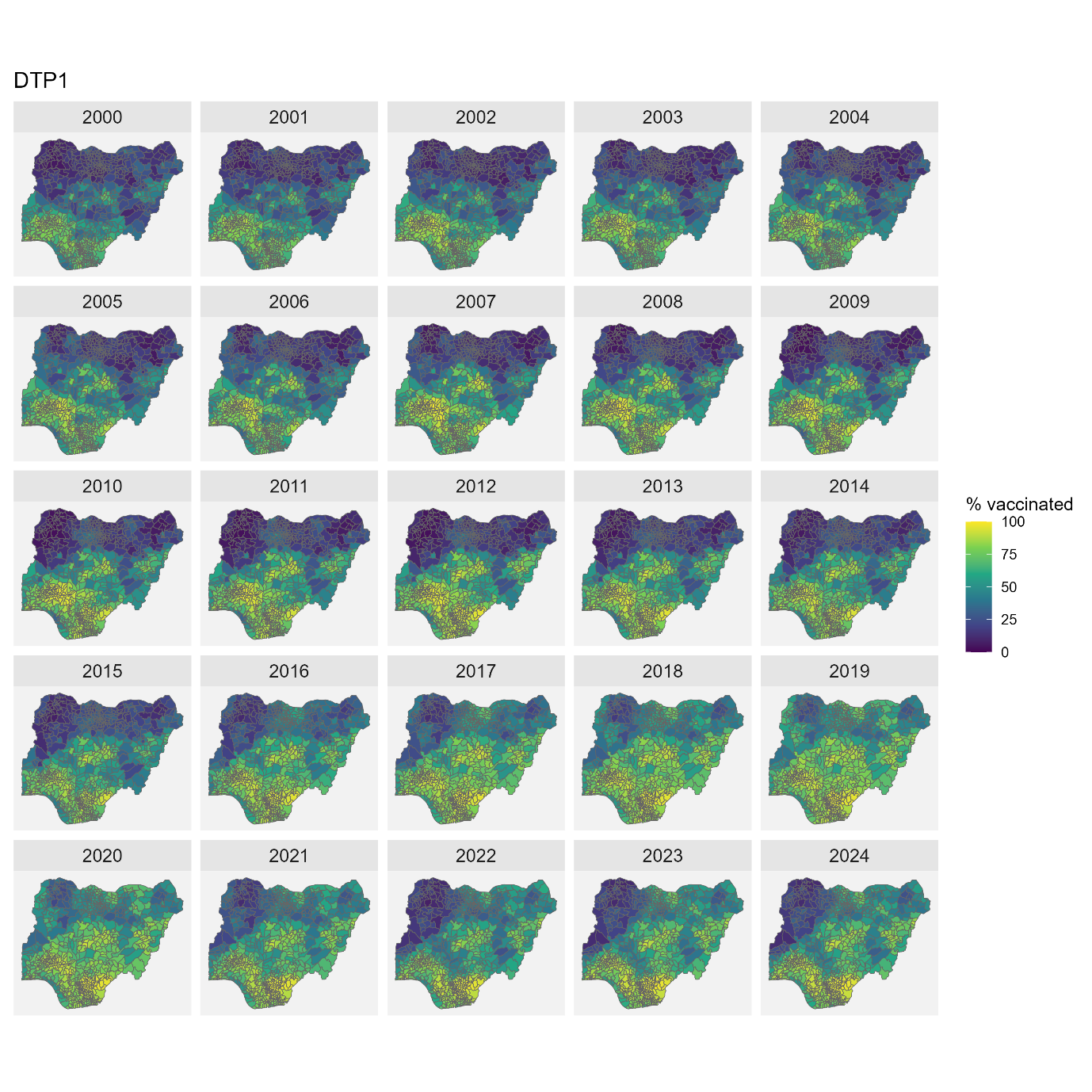

Figure 15: Distribution of vaccination coverage at the local government area level from 2000 to 2024, using DTP1 coverage as an example. These estimates relate to the birth cohort in each year. The associated uncertainties are shown in supplementary Figure 16.

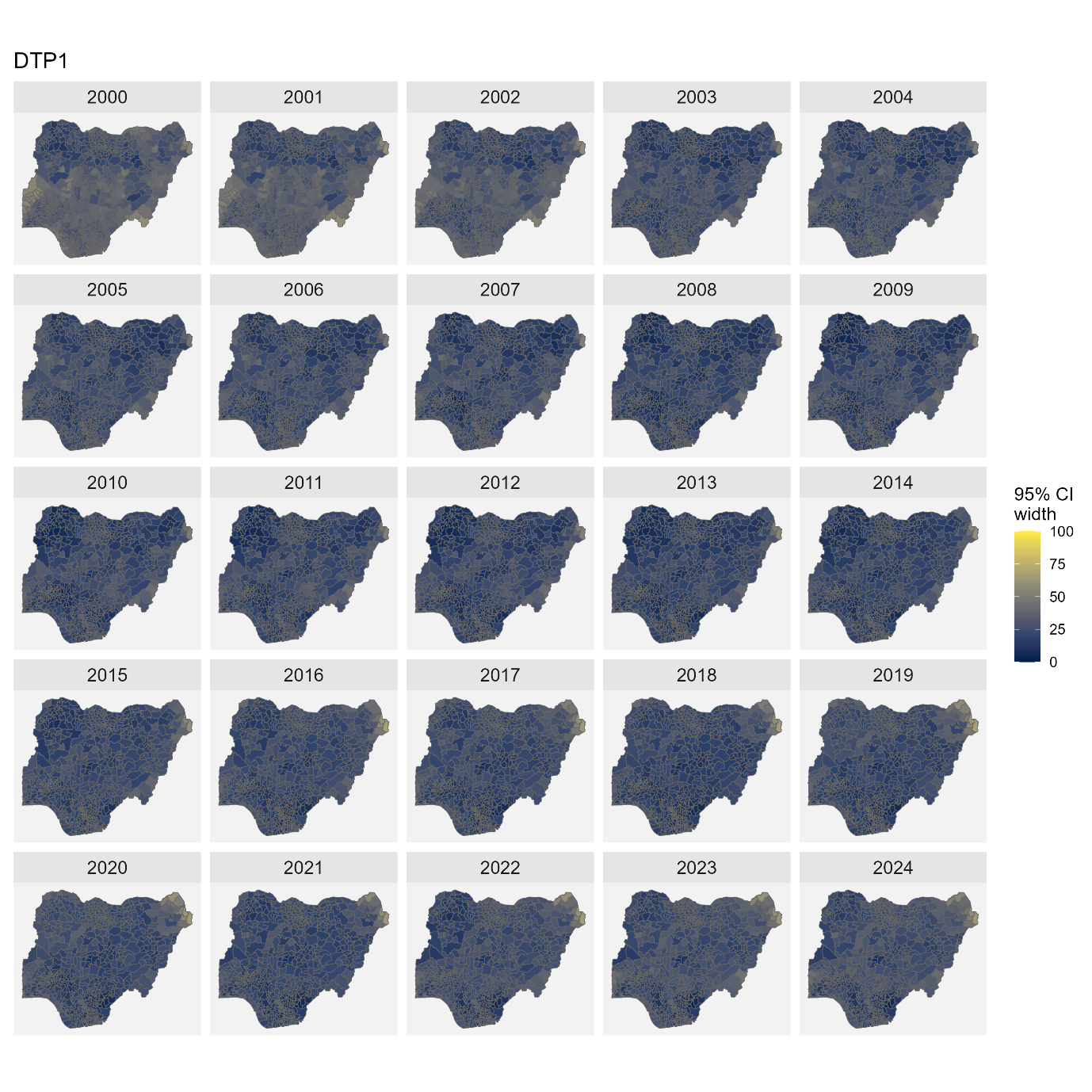

Figure 16: Local government area (LGA) level uncertainty estimates for DTP1 from 2000 to 2024, shown as the widths of the 95% credible intervals. These estimates relate to the birth cohort in each year.

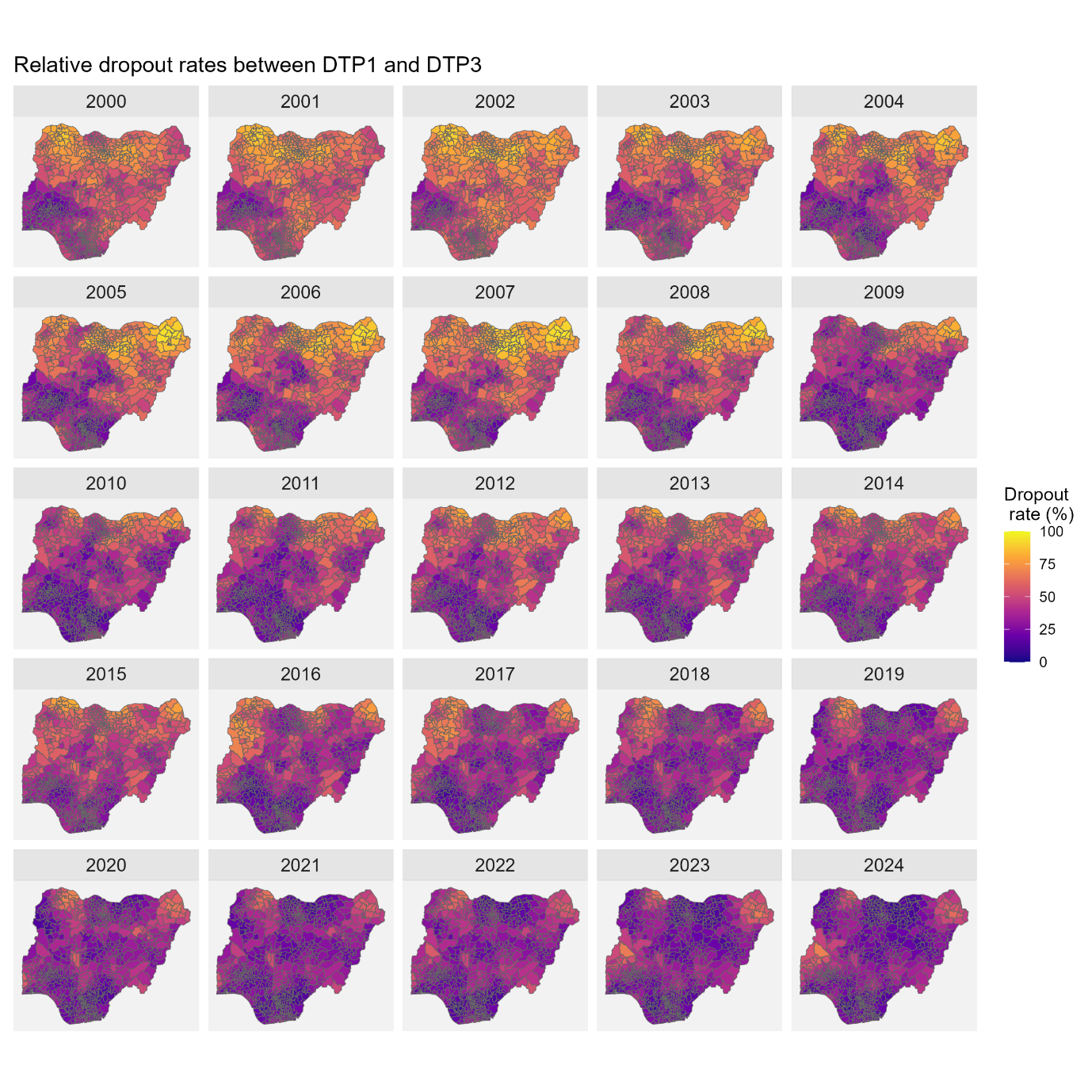

Figure 17: Dropout rates between DTP1 and DTP3 at the local government area (LGA) level.

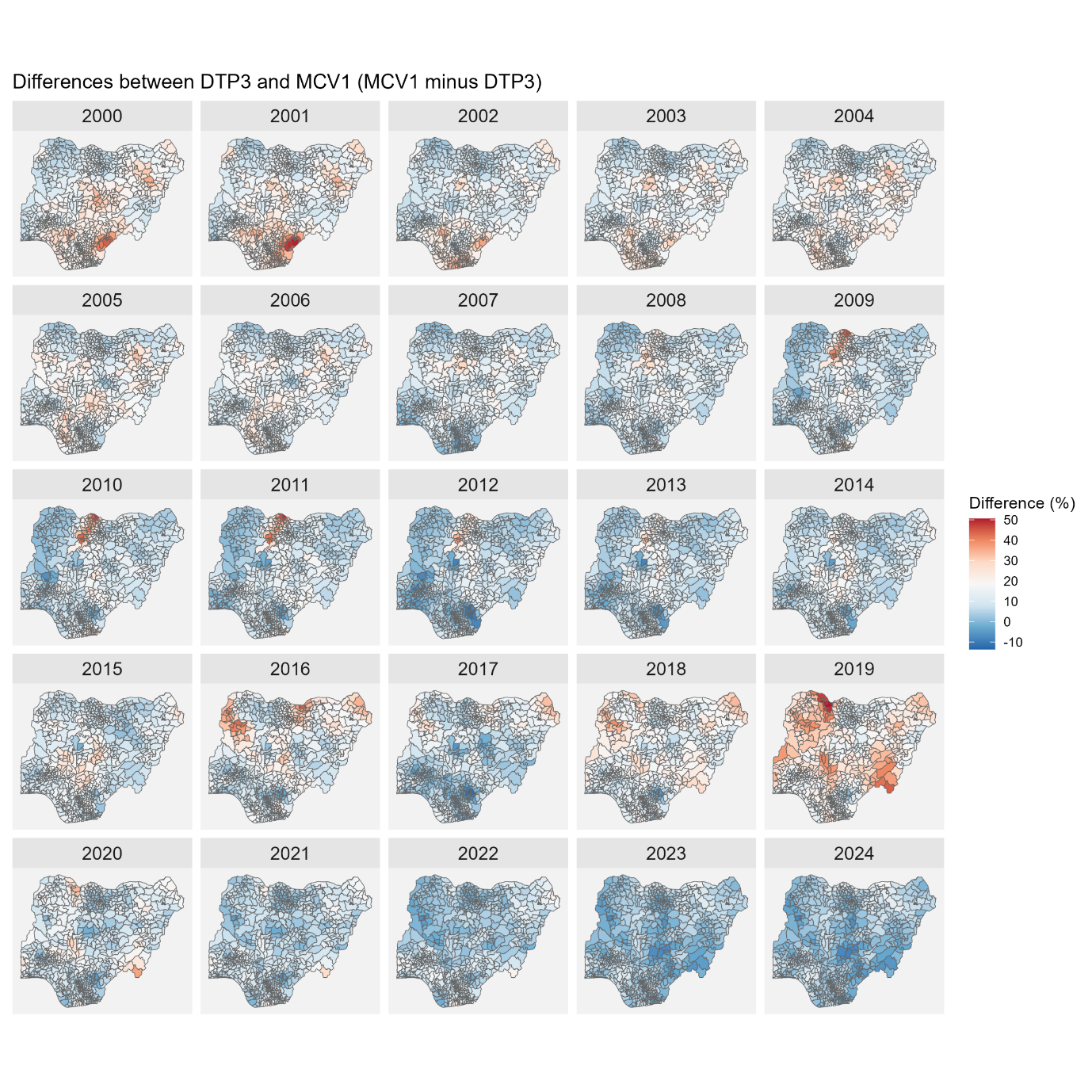

Figure 18: Differences between DTP3 and MCV1 coverage at the local government area level to investigate the effects of vaccination campaigns on MCV1 coverage.

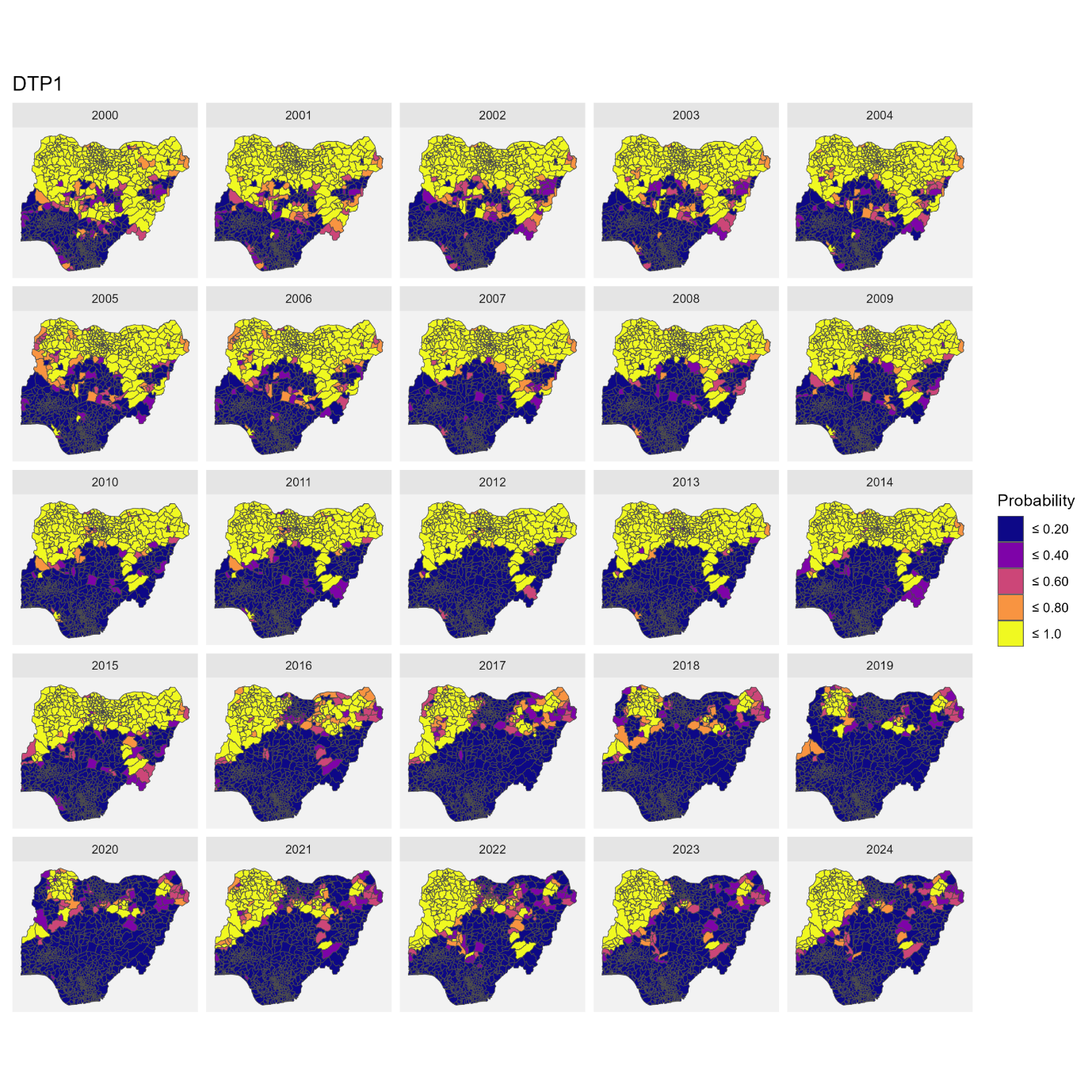

Figure 19: Probabilities of attaining an estimated DTP1 coverage of ≤40% at the local government area (LGA) level. Low coverage areas were defined as those with probabilities greater than 80%. The years shown correspond to the birth years of the cohorts included in the study.

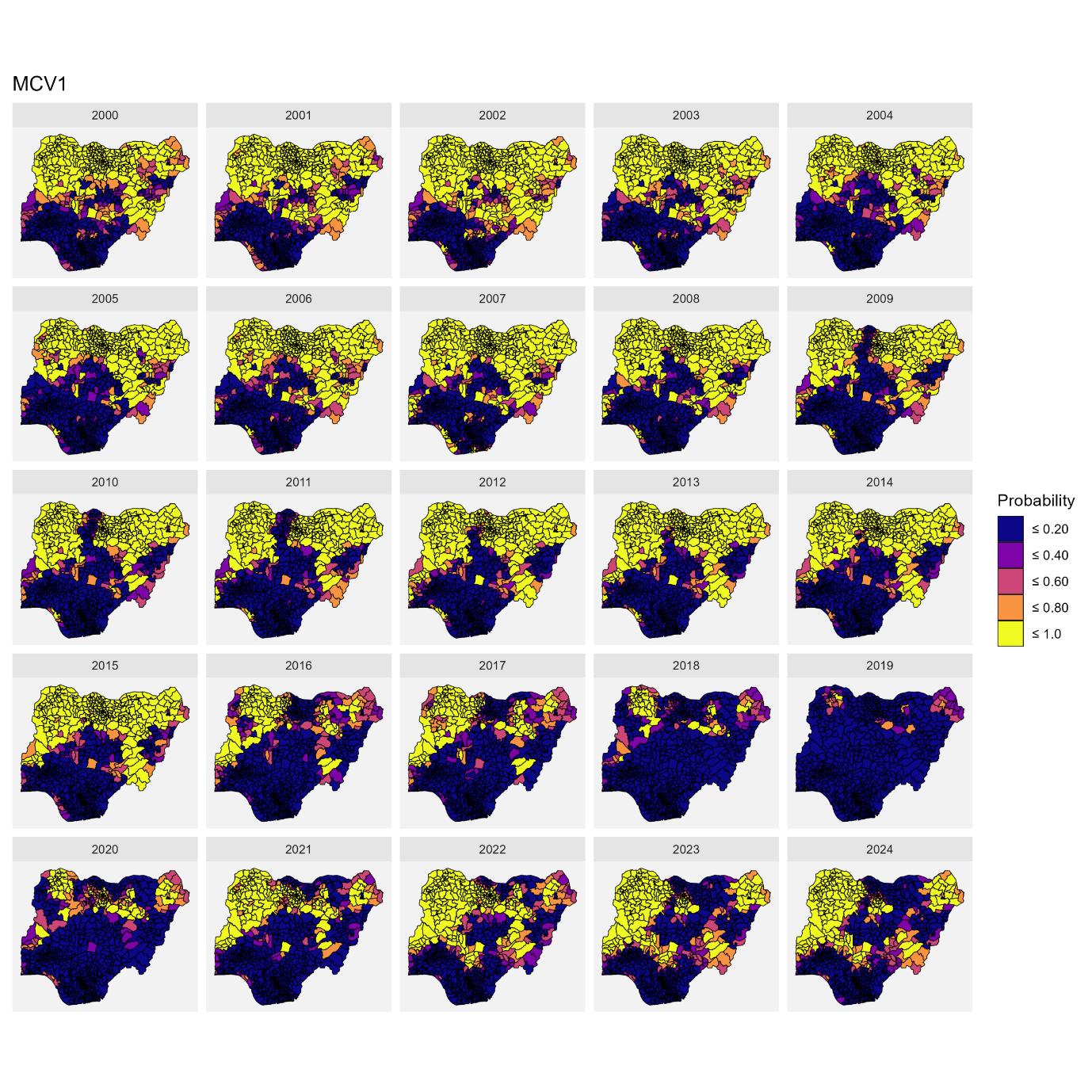

Figure 20: Probabilities of attaining an estimated MCV1 coverage of ≤40% at the local government area (LGA) level. Low coverage areas were defined as those with probabilities greater than 80%. The years shown correspond to the birth years of the cohorts included in the study.

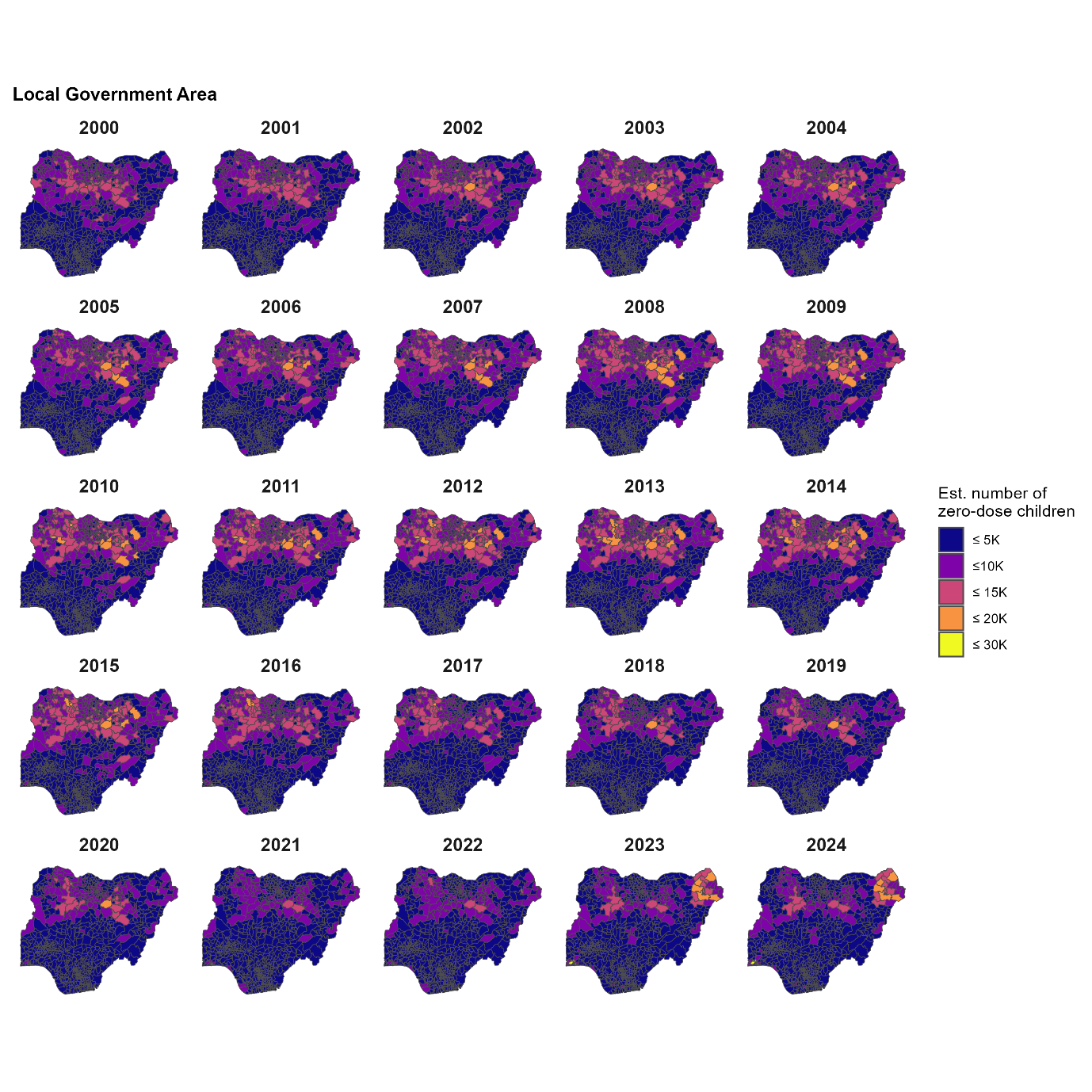

Figure 21: Estimates of numbers of (DTP) zero-dose children aged under 1 from 2000 to 2024 at the local government area level

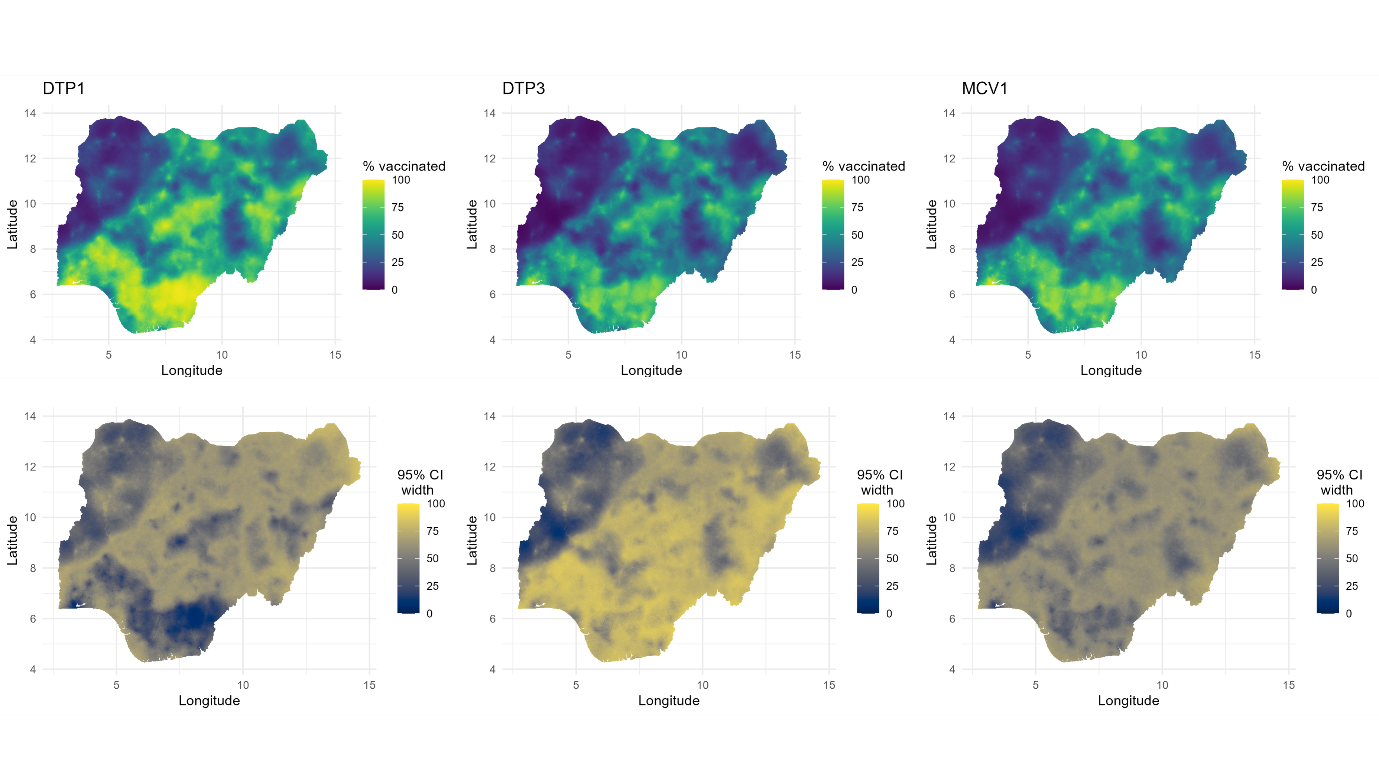

Figure 22: Predicted 1x1 km estimates of DTP1,3 and MCV1 coverage and associated uncertainties (95% credible interval width) for 2025.
